# Biomarkers Selection for Population Normalization in SARS-CoV-2 Wastewater-based Epidemiology

**DOI:** 10.1101/2022.03.14.22272359

**Authors:** Shu-Yu Hsu, Mohamed B. Bayati, Chenhui Li, Hsin-Yeh Hsieh, Anthony Belenchia, Jessica Klutts, Sally A. Zemmer, Melissa Reynolds, Elizabeth Semkiw, Hwei-Yiing Johnson, Trevor Foley, Chris G. Wieberg, Jeff Wenzel, Marc C Johnson, Chung-Ho Lin

## Abstract

Wastewater-based epidemiology (WBE) has been one of the most cost-effective approaches to track the SARS-CoV-2 levels in the communities since the COVID-19 outbreak in 2020. Normalizing SARS-CoV-2 concentrations by the population biomarkers in wastewater can be critical for interpreting the viral loads, comparing the epidemiological trends among the sewersheds, and identifying the vulnerable communities. In this study, five population biomarkers, pepper mild mottle virus (pMMoV), creatinine (CRE), 5-hydroxyindoleacetic acid (5-HIAA), caffeine (CAF) and its metabolite paraxanthine (PARA) were investigated for their utility in normalizing the SARS-CoV-2 loads through developed direct and indirect approaches. Their utility in assessing the real-time population contributing to the wastewater was also evaluated. The best performed candidate was further tested for its capacity for improving correlation between normalized SARS-CoV-2 loads and the clinical cases reported in the City of Columbia, Missouri, a university town with a constantly fluctuated population. Our results showed that, except CRE, the direct and indirect normalization approaches using biomarkers allow accounting for the changes in wastewater dilution and differences in relative human waste input over time regardless flow volume and population at any given WWTP. Among selected biomarkers, PARA is the most reliable population biomarker in determining the SARS-CoV-2 load per capita due to its high accuracy, low variability, and high temporal consistency to reflect the change in population dynamics and dilution in wastewater. It also demonstrated its excellent utility for real-time assessment of the population contributing to the wastewater. In addition, the viral loads normalized by the PARA-estimated population significantly improved the correlation (*rho*=0.5878, *p*<0.05) between SARS-CoV-2 load per capita and case numbers per capita. This chemical biomarker offers an excellent alternative to the currently CDC-recommended pMMoV genetic biomarker to help us understand the size, distribution, and dynamics of local populations for forecasting the prevalence of SARS-CoV2 within each sewershed.

**HIGHLIGHT (bullet points):**

1. The paraxanthine (PARA), the metabolite of the caffeine, is a more reliable population biomarker in SARS-CoV-2 wastewater-based epidemiology studies than the currently recommended pMMoV genetic marker.
2. SARS-CoV-2 load per capita could be directly normalized using the regression functions derived from correlation between paraxanthine and population without flowrate and population data.
3. Normalizing SARS-CoV-2 levels with the chemical marker PARA significantly improved the correlation between viral loads per capita and case numbers per capita.
4. The chemical marker PARA demonstrated its excellent utility for real-time assessment of the population contributing to the wastewater.

## 1. INTRODUCTION

Severe acute respiratory syndrome coronavirus 2 (SARS-CoV-2), has caused a pandemic declared by the World Health Organization (WHO) on March 11th, 2020 [1]. Despite clinical tests being sufficient and accurate, their time-consuming and often expensive process has not always been sufficient enough to track SARS-CoV-2 outbreaks at the population scale[2]. Wastewater-based epidemiology (WBE) offers near real-time information about the outbreak to track SARS-CoV-2 in the communities [3]. It has been successfully used to predict the overall status of infection and to capture asymptomatic and pre-symptomatic infections in the given wastewater treatment plant (WWTP) served area [4]. Several studies in Europe, Australia, Japan, Singapore and the United States had used WBE approach. [4–12]. The State of Missouri launched a statewide wastewater SARS-CoV-2 surveillance program in May 2020. [13]. It has been successfully applied to 1) provide the early warning, 2) determine the distribution of SARS-CoV-2 and its variants in Missouri, 3) identify trends in SARS-CoV-2 prevalence in areas surveilled, and 4) monitor for indicators of SARS-CoV-2 reemergence to inform mitigation efforts.

For long-term wastewater SARS-CoV-2 surveillance, normalizing SARS-CoV-2 wastewater concentrations prior to calculating trends is recommended by the United States Centers for Disease Control (CDC) to account for changes in wastewater dilution and differences in relative human waste input over time, due to tourism, weekday commuters, temporary workers, etc. Normalizing SARS-CoV-2 concentrations by the amount of human feces in wastewater can be crucial for interpreting and comparing viral concentrations in the sewage samples over time [14].

The recommended population biomarkers include organisms or chemical compounds specific to human feces that can be measured in wastewater to estimate the size of the population. These biomarkers include but are not limited to viral or bacterial molecular targets [15]. Pepper Mild Mottle Virus (pMMoV), a viral pathogen in *Capsicum sp*. that had been identified in several pepper-based products and diets [16], is one of the biomarkers recommended by the CDC [17]. Due to the abundance in pepper-based food, unaffected by seasonal change, persistence in the wastewater (with half-life from 6-10 days) from the populated area, the pMMoV was recognized as one of the promising population biomarkers [18,19].

In addition to the viral or bacterial genetic markers, small chemical molecules biomarkers were also utilized to estimate the population at the area served by given WWTP [20–25]. Several chemical markers, such as creatinine (CRE), 5-hydroxyindoleacetic acid (5-HIAA), caffeine (CAF), and its metabolite paraxanthine (PARA) have been reported as promising candidates [20–25]. Creatinine is the metabolite of creatine and phosphorylcreatine in the muscles. It is produced at a steady state, diffused out of muscle cells, and further excreted by kidneys into urine [26]. Urinary CRE was routinely used to account for dilution when testing human urine for illicit substances [27,28]. The serotonin metabolite 5-hydroxyindoleacetic acid (5-HIAA) is the other promising endogenous molecule for this purpose. Clinical urinary 5-HIAA analysis is commonly performed to evaluate patients with suspected carcinoid syndrome [29]. The 5-HIAA in the wastewater was also used to estimate the population [22]. Both CRE and 5-HIAA had been quantified in the samples from WWTPs [30,31]. Rico *et al*. reported that 5-HIAA loads in the WWTP samples showed a positive correlation with the population calculated using the hydrochemical parameters [22]. Chen *et al*. reported that 5-HIAA levels were also correlated well with the census population [23].

In additional to endogenous molecules, CAF, a widely consumed central nervous system (CNS) stimulant [32], is commonly found in food products, including tea, coffee, and energy drinks, as well as in some medications and dietary supplements. The PARA is the major metabolite of CAF through the cytochrome P4501A2 (CYP1A2)-catalyzed 3-demethylation[33]. Several studies had detected CAF and PARA in the wastewater [21,24,25,30,34]. Similar to 5-HIAA, researches have reported a positive correlation between CAF load and the population from census or population calculated by the hydrochemical parameters [21,22]. The PARA level was found less affected by the genetic heterogeneity and population structure as compared to its parent compound CAF [33], suggesting PARA could also be a potential population biomarker.

The goal of this study was to determine the most suitable population biomarker for SRAS-CoV-2 wastewater surveillance. Specific objectives were 1) to compare the variability and accuracy of the selected biomarkers for normalizing the SARS-CoV2 concentrations using two different approaches, 2) to identify the suitable biomarkers for estimating the real-time population contributing to the wastewater, and 3) to demonstrate the normalized SARS-CoV-2 loads per capita with the selected biomarkers against the clinic cases.

## 2. MATERIAL AND METHOD

### 2.1 Chemicals and reagents

All of the analytical standards were purchased from Sigma-Aldrich (St. Louis, MO, USA) except 5-Hydroxyindoleacetic acid-[13C6] (5-HIAA-[13 C6]) (≥98%) was purchased from IsoSciences (Ambler, PA, USA). The HPLC grade methanol and acetonitrile used in these experiments were purchased from Sigma-Aldrich (St. Louis, MO, USA). The TaqPath™ 1-Step RT-qPCR Master Mix and the TaqMan probe for *pMMoV* gene detection were purchased from Fisher Scientific (USA). The primers and the TaqMan probes for N1 and N2 gene detections were purchased from Integrated DNA Technologies, Inc. (USA). Waters Oasis HLB SPE cartridge (500 mg) was purchased from Waters Milford, MA (USA). Whatman® Anotop® filters were purchased from Fisher Scientifics (USA).

### 2.2 Wastewater sampling

To develop the relationship between biomarkers and population, triplicates of 50 mL of the 24-hour composite wastewater samples were collected once per week from the raw inlets, before the primary treatment, at 12 WWTPs (Table 1) in Missouri from 18th to 29th in January 2021. Following the correlation analysis, wastewater composite samples collected from 64 WWTPs (Table S1) across the State of Missouri, were used for method validation. They were collected during the week of May 10th in 2021. The WWTPs serve urban, semirural, and rural locations throughout Missouri with the sewershed population ranging from 4,600 to 306,647 (number of people estimated by WWTPs or Missouri Census). Ten wastewater composite samples were collected from WWTPs at the City of Columbia (college town) and a tourist town respectively through May to early September in 2021 (Table S2) for evaluating the utility of the biomarker for assessing the population fluctuation and dynamics. All of wastewater samples were transported in coolers with cold packs and then stored at 4°C until further extraction within two days.

**Table 1.**
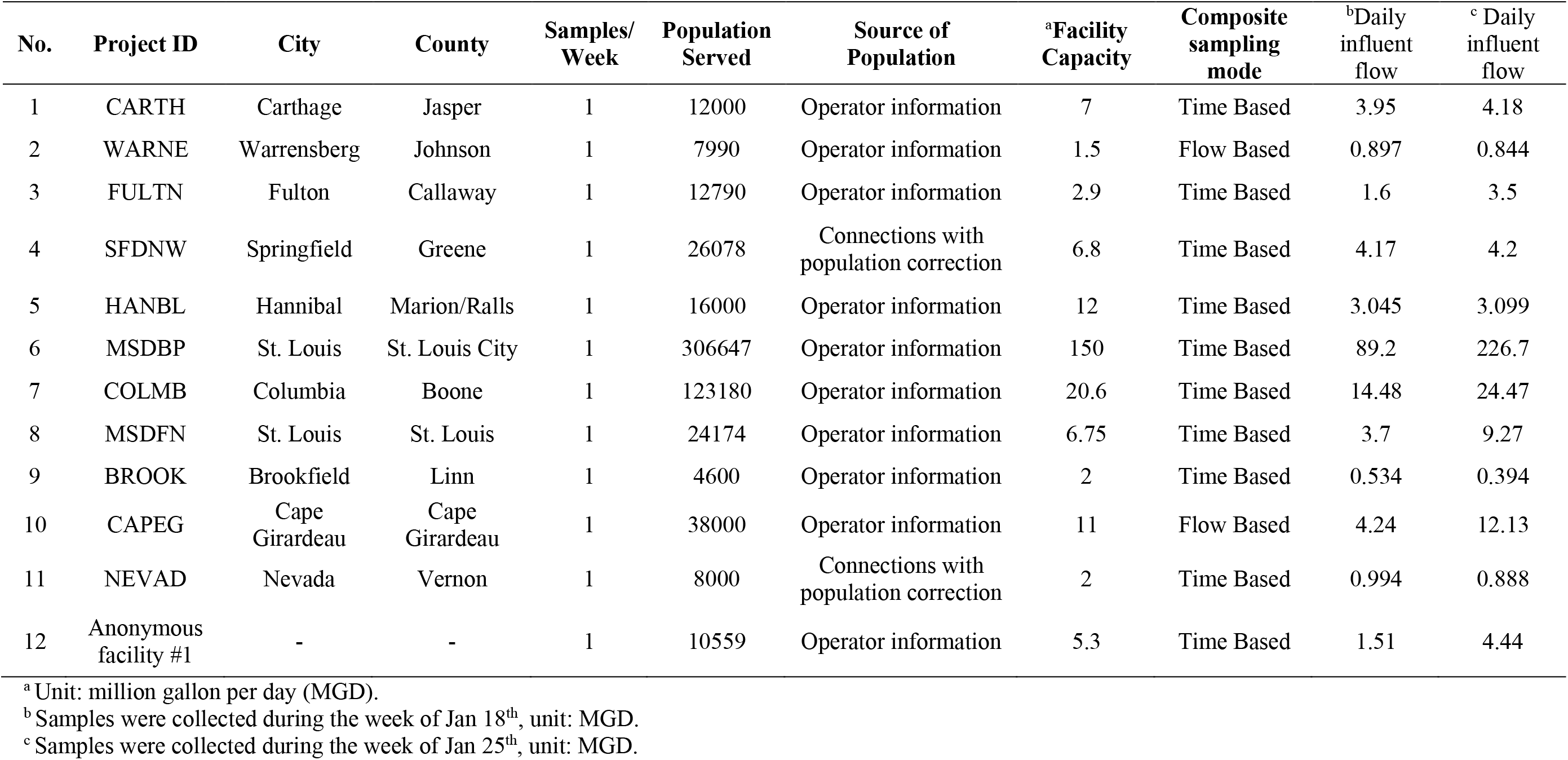
Site summary of the 12 wastewater treatment plants for the model development.

| No. | Project ID | City | County | Samples/<br>Week | Population<br>Served | Source of<br>Population | <sup>a</sup>Facility<br>Capacity | Composite<br>sampling<br>mode | <sup>b</sup>Daily<br>influent<br>flow | <sup>c</sup> Daily<br>influent<br>flow |
| --- | --- | --- | --- | --- | --- | --- | --- | --- | --- | --- |
| 1 | CARTH | Carthage | Jasper | 1 | 12000 | Operator information | 7 | Time Based | 3.95 | 4.18 |
| 2 | WARNE | Warrensberg | Johnson | 1 | 7990 | Operator information | 1.5 | Flow Based | 0.897 | 0.844 |
| 3 | FULTN | Fulton | Callaway | 1 | 12790 | Operator information | 2.9 | Time Based | 1.6 | 3.5 |
| 4 | SFDNW | Springfield | Greene | 1 | 26078 | Connections with<br>population correction | 6.8 | Time Based | 4.17 | 4.2 |
| 5 | HANBL | Hannibal | Marion/Ralls | 1 | 16000 | Operator information | 12 | Time Based | 3.045 | 3.099 |
| 6 | MSDBP | St. Louis | St. Louis City | 1 | 306647 | Operator information | 150 | Time Based | 89.2 | 226.7 |
| 7 | COLMB | Columbia | Boone | 1 | 123180 | Operator information | 20.6 | Time Based | 14.48 | 24.47 |
| 8 | MSDFN | St. Louis | St. Louis | 1 | 24174 | Operator information | 6.75 | Time Based | 3.7 | 9.27 |
| 9 | BROOK | Brookfield | Linn | 1 | 4600 | Operator information | 2 | Time Based | 0.534 | 0.394 |
| 10 | CAPEG | Cape<br>Girardeau | Cape<br>Girardeau | 1 | 38000 | Operator information | 11 | Flow Based | 4.24 | 12.13 |
| 11 | NEVAD | Nevada | Vernon | 1 | 8000 | Connections with<br>population correction | 2 | Time Based | 0.994 | 0.888 |
| 12 | Anonymous<br>facility #1 | - | - | 1 | 10559 | Operator information | 5.3 | Time Based | 1.51 | 4.44 |

<sup>a</sup> Unit: million gallon per day (MGD).
<sup>b</sup> Samples were collected during the week of Jan 18<sup>th</sup>, unit: MGD.
<sup>c</sup> Samples were collected during the week of Jan 25<sup>th</sup>, unit: MGD.

### 2.3 Detection of SARS-CoV-2 concentration

#### 2.3.1 RNA extraction from wastewater samples

Fifty mL of wastewater from each catchment was filtered through a 0.22-micron filter (Millipore cat# SCGPOO525). Thirty-six mL of filtered wastewater were mixed with 12 mL of 50% (W/V) polyethylene glycol (PEG, Research Products International, cat# P48080) and 1.2 M NaCl, followed by incubation for 2 hours at 4°C. Samples were further centrifuged at 12,000 Xg for 2 hours. RNA was extracted from the pellet using Qiagen Viral RNA extraction kit following the manufacturer’s instructions after the supernatant was removed. RNA was eluted in a final volume of 60 μL. The samples were stored at -80°C if they couldn’t be processed immediately.

#### 2.3.2 Plasmid standard preparation

A plasmid carrying a *pMMoV* gene 180-bp fragment (Table 2) along with a N gene fragment was constructed, purified from *Escherichia coli*, and used as standards for the RT-qPCR assay. The primer pair, COVID19-N 5p and COVID19-N 3p (Table 2), was used to amplify the N ORF fragment from IDT’s 2019-nCoV_N_Positive Control plasmid and the N ORF fragments were infused using an InFusion kit (Takara) as described [35]. A standard curve was constructed at concentrations of 200,000 through 2 gene copies μL^-1^ and utilized to determine the copy number of the target *pMMoV* gene in the spiked wastewater samples.

**Table 2.**
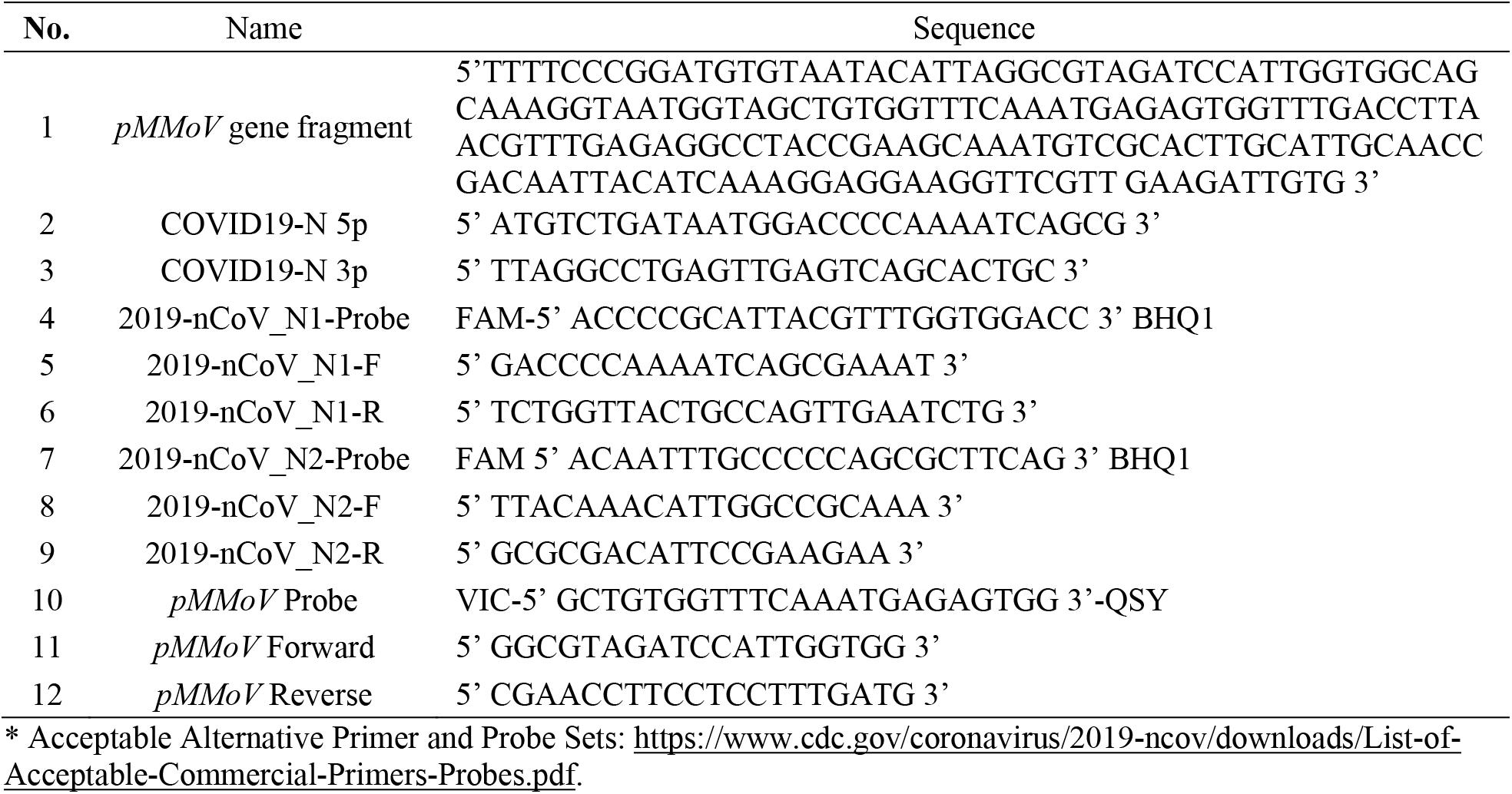
The sequences of *pMMoV*, primers, and probes.

#### 2.3.3 Quantitative RT-qPCR assay

The TaqMan probe 2019-nCoV_N1-Probe and the primer pair (2019-nCoV_N1-F and 2019-nCoV_N1-R) for N1 detection, and The TaqMan probe 2019-nCoV_N2-Probe and the primer pair (2019-nCoV_N2-F and 2019-nCoV_N2-R) for N2 detection from Integrated DNA Technologies (IDT) were chosen based on the CDC 2019-nCoV Real-Time RT-PCR Diagnostic Panel (Acceptable Alternative Primer and Probe Sets). The sequences of probes and primers were listed in Table 2. Final RT-qPCR one-step mixtures for *N1/N2* or *pMMoV* detection consisted of 5 μL TaqPath 1-step RT-qPCR Master Mix (Thermo Fisher), 500 nM of each primer, 125 nM of the TaqMan probes, 5 μl of wastewater RNA extract, and RNase/DNase-free water to reach a final volume of 20 μL. All RT-qPCR assays were performed in duplicate using a 7500 Fast real-time qPCR System (Applied Biosystems). The reactions were initiated with 1 cycle of UNG incubation at 25°C for 2 min and then 1 cycle of reverse transcription at 50°C for 15 min, followed by 1 cycle of activation of DNA polymerase at 95°C for 2 min and then 45 cycles of 95°C for 3 sec for DNA denaturation and 55°C for 30 sec for annealing and extension. The data would be collected at the step of 55°C extension.

### 2.4 Quantification of biomarkers

#### 2.4.1 Detection of pMMoV viral concentration

The TaqMan probe (*pMMoV* Probe) and the primer pair (*pMMoV* Forward and *pMMoV* Reverse, Table 2) were designed and used to target the *pMMoV* RNA. The specificity of primers and probe were tested by BLAST analysis (NCBI) to prevent known nonspecific binding targets that could be obtained in a human specimen. The *pMMoV* concentration in the wastewater sample is determined by the quantitative RT-qPCR assay as described above.

#### 2.4.2 Extraction of 5-hydroxyindoleacetic acid

The wastewater was filtered through a 0.2 μm Whatman® Anotop® filter. Twenty ml of filtered wastewater was fortified with 20 μL of 100 ppm 5-HIAA-^13^C6 followed by solid-phase extraction (SPE) using Waters Oasis HLB SPE cartridge (500 mg). The extracts on the SPE cartridge were eluted with the mixture of 50% acetonitrile (ACN) and 50% methanol. The samples were resuspended with ACN after evaporation. Samples were stored at -20 °C until analyzed by the high-performance liquid chromatography-tandem mass spectrometry (LC-MSMS) analysis.

#### 2.4.3 Extraction of creatinine, caffeine, and paraxanthine

One thousand and six hundred μL of a subsample from filtered wastewater was spiked with 10 μL of formic acid followed by a vortexing vigorously. The mixture was centrifuged at 10,000 rpm for 10 mins. Seven hundred fifty μl of supernatant was mixed with 750 μl of LC-MSMS buffer (10 mM ammonium acetate and 0.1% formic acid in water) followed by fortification of 20 μl of 76 ppm caffeine-C^13^ or creatinine-D_3_. The mixture was filtered through a 0.2 μm Anotop PTFE filter before the LC-MSMS analysis.

#### 2.4.4 Liquid chromatography-tandem mass spectrometry analysis

The quantification of 5-HIAA, creatinine, caffeine, and paraxanthine was performed by a Waters Alliance 2695 High Performance Liquid Chromatography (HPLC) system coupled with Waters Acquity TQ triple quadrupole mass spectrometer (MS/MS). The analytes were separated using a Phenomenex (Torrance, CA) Kinetex C18 (100mm x 4.6 mm; 2.6 μm particle size) reverse-phase column. The mobile phase consisted of (A)10 mM ammonium acetate and 0.1% formic acid in water and (B) 100% acetonitrile. The gradient conditions were 0 – 0.3 min, 2% B; 0.3-7.27 min, 2-80% B; 7.27-7.37 min, 80-98% B; 7.37-9.0 min, 98% B; 9-10 min 98-2% B; 10.0 – 15.0 min, 2% B at the flow rate of 0.5 mL/min. The ion source in the MS/MS system was electrospray ionization (EI) operated in either positive or negative ion mode with a capillary voltage of 1.5 kV. The ionization sources were programmed at 150°C and the desolvation temperature was programmed at 450°C. The optimized collision energy, cone voltage, molecular and product ions of biomarkers are summarized in Table 3.

**Table 3.**
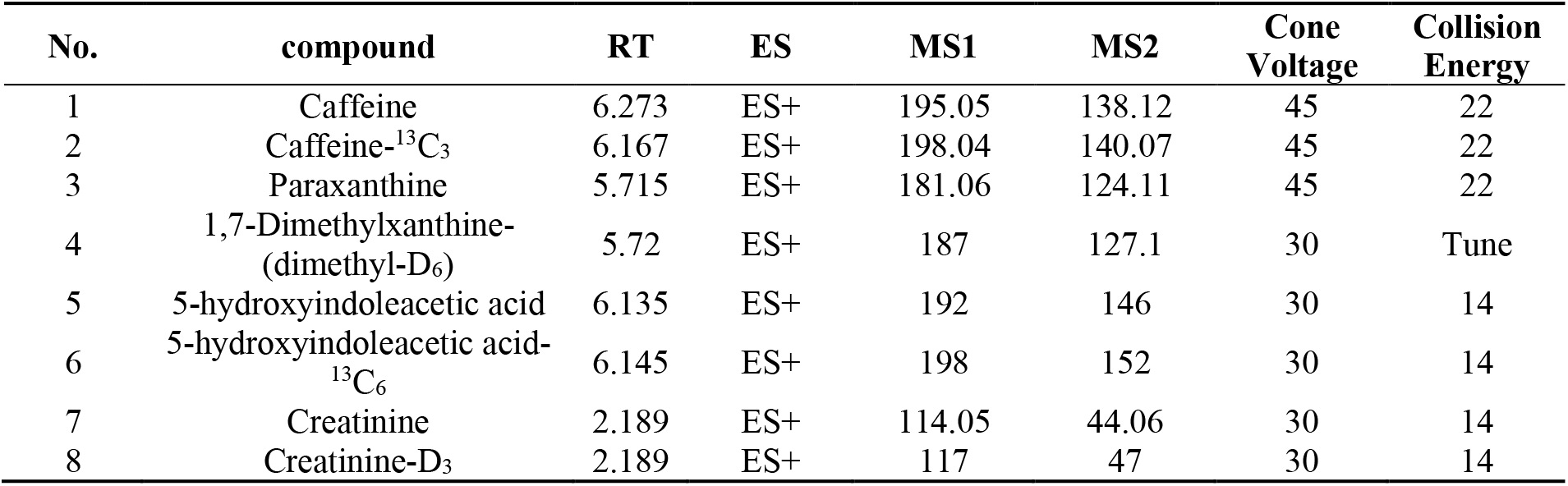
Summary of the optimized LC-MSMS Parameters for chemical population biomarkers.

### 2.5 Normalization of SARS-CoV-2 concentration with biomarker concentration

Two approaches were proposed to normalize SARS-CoV-2 concentration in the wastewater using the established regression functions from the linear regression models, assuming that the biomarker load is proportional to the population in the wastewater composite (Fig. 1). This section presents the methods of (1) determining the regression functions and (2) normalizing SARS-CoV-2 concentrations using biomarker concentrations are presented.

**Figure 1.**
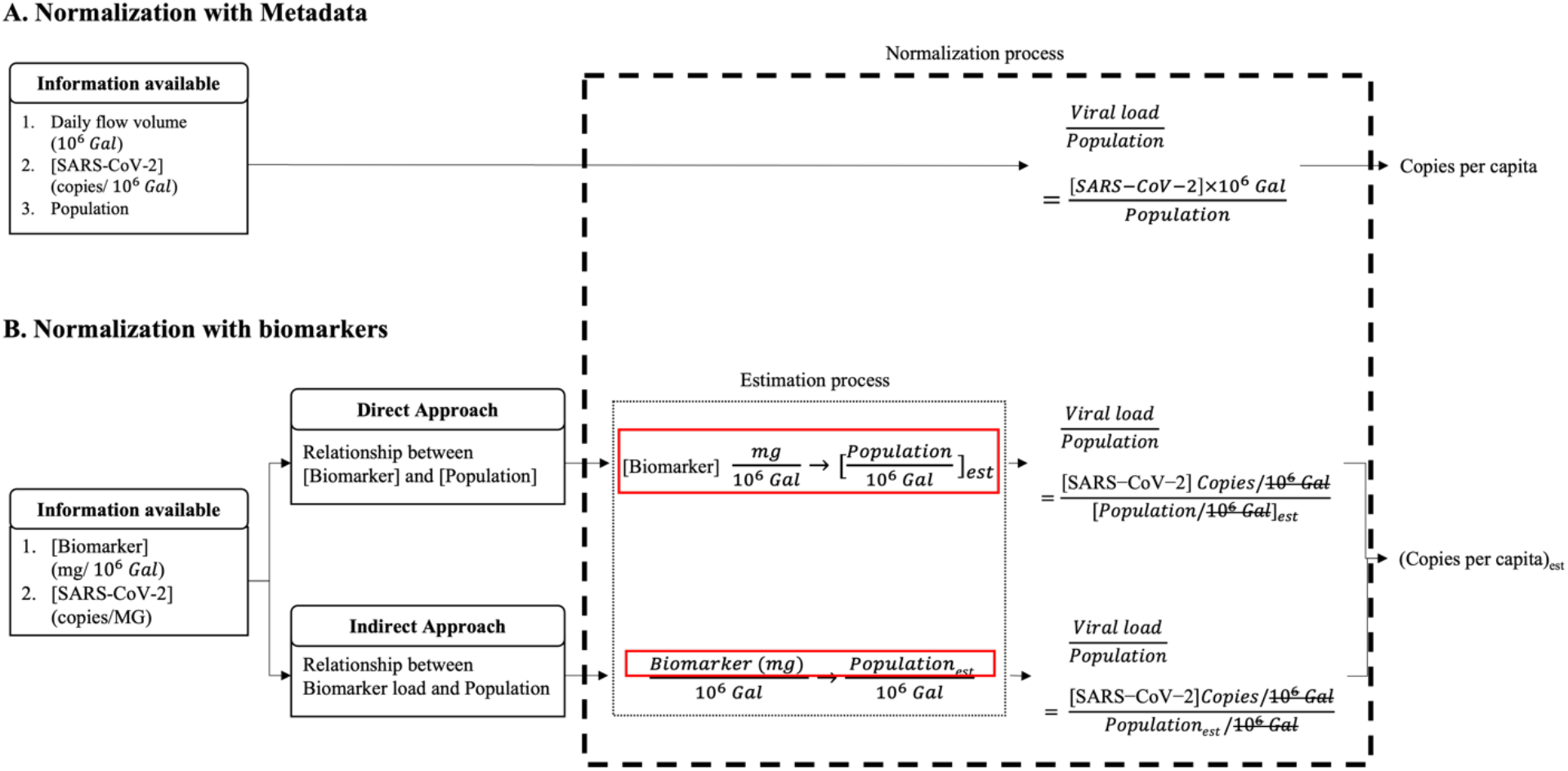
Normalization processes of determining SARS-CoV-2 load per capita. (A) When the population size, daily flow volume and viral concentration of the metadata are used in the normalization process. (B) When the real-time population size of the sewershed is estimated using regression functions developed from the correlation between biomarker and population size from metadata in direct or indirect approach.

#### 2.5.1 Relationships between biomarker concentration and population concentration in wastewater

The population concentration is expressed as

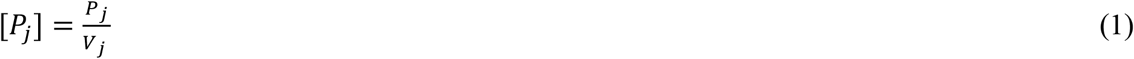

in which, [*P* _*j*_] is the population concentration in the wastewater for WWTP *j*. Both the population *P*_*j*_ and the daily flow volume *V*_*j*_ (MGal, million gallons) for WWTP *j* are provided in metadata (Table 1). The population concentration [*P*] is modeled as

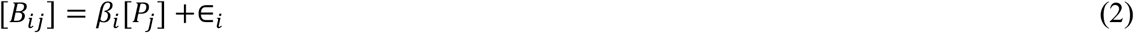

where [*B* _*ij*_] is the concentration of biomarker *i* in WWTP *j* sample, the corresponding population concentration [*P* _*j*_], the error term *∈*_*i*_, and the estimated parameter *β*_*i*_ for biomarker *i*. The error term accounts for differences in biomarker concentration from daily variations at the locations. To avoid any skewness, Log-transformed population and biomarker concentrations were further used to fit a linear regression model. The Pearson’s correlation coefficient (*r*) was calculated.

#### 2.5.2 Relationships between biomarker loads and population size

Daily flow volume was taken into consideration before the relationship between daily biomarker load and the population contributing to the wastewater was examined. The biomarker load of biomarker *i* for WWTP *j, B*_*ij*_, was calculated as

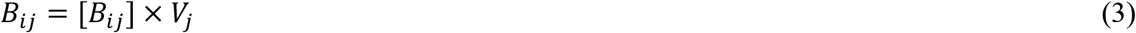

in which, [*B* _*ij*_], the biomarker *i* concentration in WWTP *j* wastewater samples, was determined by LC-MSMS. The population *P* is modeled as

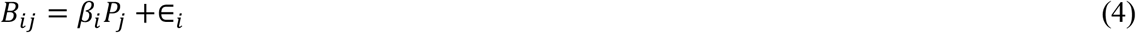

Where *B*_*ij*_ is the daily *i* biomarker load, *P*_*j*_ the population from metadata at WWTP *j*.

#### 2.5.3 Developing the normalization scheme derived from metadata

According to the CDC’s guideline, the normalization of SARS-CoV-2 load (copy/person/day) is expressed and calculated as

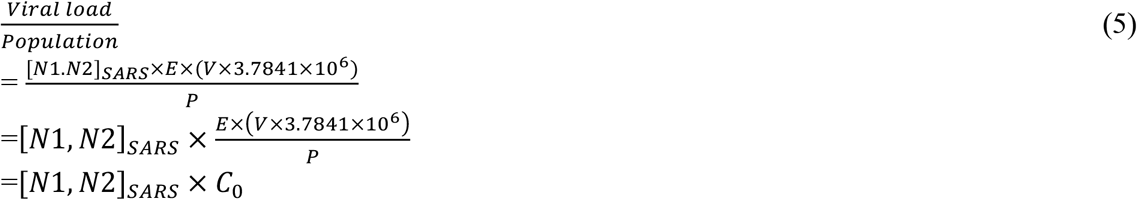

in which, *[N1, N2]*_*SARS*_ (copies/μL) is the average of replicated N1 and N2 concentrations (n=4) in the wastewater samples. *E*, concentration factor, 350, transforms unit of concentration from copies/μL of RNA to copies/L of wastewater. Daily flow volume *V* ‘(MGal, million gallons) and population *P* are provided in Metadata. A constant, 3.78541, is applied to convert the imperial unit to metric unit. In the las line, all variables and constants are designated as normalization coefficient 0 (*C*_*0*_) except [*N1,N2]*_*SARS*_. The unit of normalized SARS-CoV-2 load per capita turns into copies per person.

#### 2.5.4 Developing the normalization scheme derived from the relationship between biomarker concentration and population concentration

The population concentration estimated by biomarker concentration in the wastewater was utilized in the ***direct*** normalization approach. The correlation between the biomarker *i* concentrations and population in wastewater is expressed as

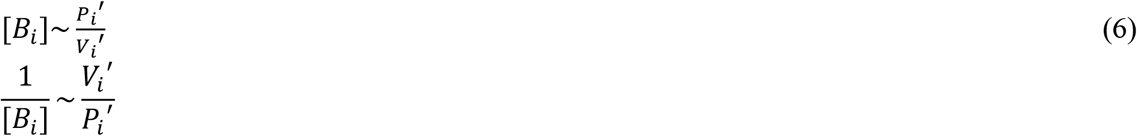

in which population *P* _*i*_*’* and daily flow volume *V* _*i*_ *‘* were estimated using biomarker *i* concentration in the Eq. (2). The reciprocal of the estimated population *P* _*i*_*’* and daily flow volume *V* _*i*_ *‘* were unitized in SARS-CoV-2 load normalization process:

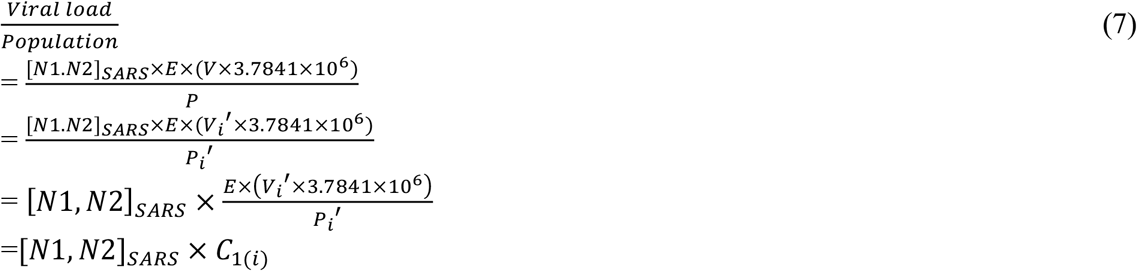

in which, the *P* and *V* in line 2 are replaced with *P’*_*i*_ and *V’*_*i*_ in Eq. (6) resulting in line 3. Except [*N1,N2]*_*SARS*_, all of variables and constants were designated as normalization coefficient 1, *C*_*1(i)*_, for biomarker *i* in the direct approach. The *C*_*1(i)*_ was further standardized by *C*_*0*_ as

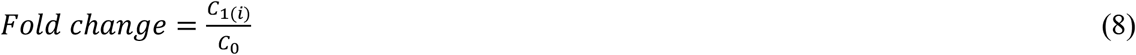

The fold change was utilized to assess the fitness, precision, and the variability of the biomarkers.

#### 2.5.5 Developing the normalization scheme derived from the relationship between biomarker loads and population

The population estimated by biomarker loads in the wastewater were used in the ***indirect*** biomarker to fall into the linear range of the correlation:

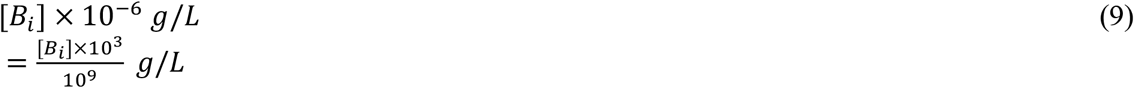

in which, [*B* _*i*_] is the concentration of biomarker *i* (μg/L or copies/L). The population was estimated using *[B]*’ 10^3^ as B in the Eq. (4), and the unit of estimated population concentration ([*P’*]) became person/L. The population concentration ([*P* _*i*_]*’*) estimated by biomarker *i* is further utilized in SARS-CoV-2 load normalization below:

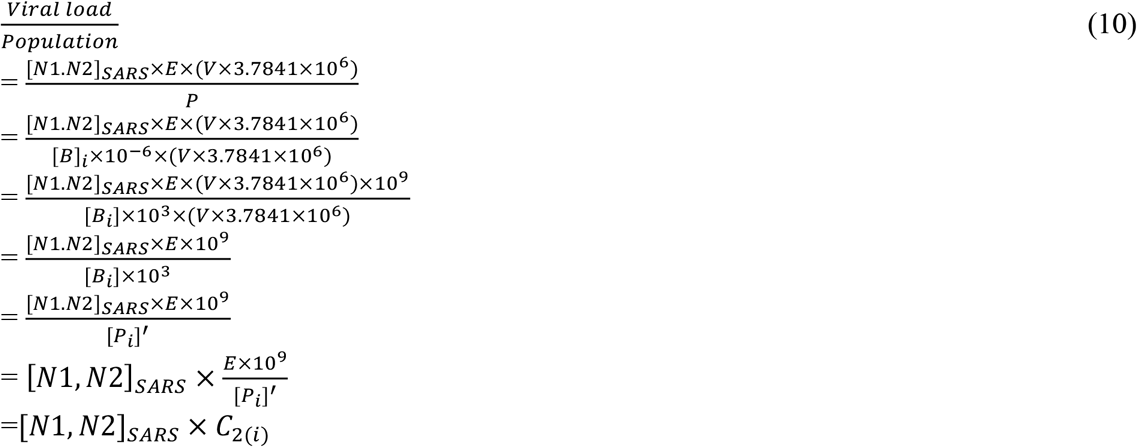

in which, the daily flow volume and constants in both numerator and denominator were canceled out in line 3, which resulted in line 4. Except [*N1,N2]*_*SARS*_, all of variables and constants were designated as normalization coefficient 2, *C*_*2(i)*_, for biomarker *i* in the indirect approach. The *C*_*2(i)*_ was further standardized by the *C*_*0*_ as

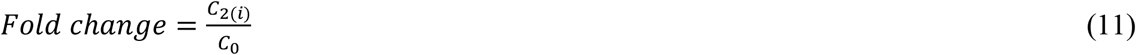

### 2.6 Validation of normalization coefficients

The regression function of two approaches were established to normalize SARS-CoV-2 load using the 24 samples collected in January 2021 (Table 1). Samples collected from 64 WWTPs in May 2021 (Table S1) were utilized as testing data set to validate the estimation of the normalization coefficients (*C*_*1(i)*_ and *C*_*2(i)*_) from two approaches. During the validation, *C*_*0*_ was calculated using Metadata in Eq. (5). The *C*_*1(i)*_ and *C*_*2(i)*_ were calculated using the concentration of CAF and PARA with Eq. (7) and (10), respectively, followed by standardization with *C*_*0*_ to evaluate the fitness, precision, and the variability.

### 2.7 Estimation of population contributing to the wastewater

#### 2.7.1 Linear regression model

To determine the accuracy and precision of population estimated by different biomarkers, the log-transformed biomarkers loads (n=24), collected from 12 WWTPs across the State of Missouri (Table1), were used as predictor variable to fit the linear regression model in R.

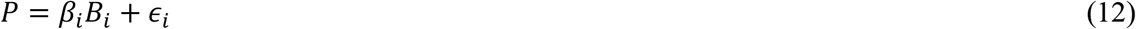

Nineteen of the data points (approximately 80%) was randomly selected as training data set to fit the model, and the rest 5 data points were used as test data set. The adjusted R^2^ and the mean square error (MSE) were utilized to evaluate the model fitting and prediction accuracy, respectively. A *k*-fold cross-validation (*k* = 5) was performed to eliminate the poor prediction from the outliers and determine the overall predictive capability of the model based the 5-fold cross-validation MSE [36].

#### 2.7.2 Estimation of real-time populations for City of Columbia (college Town) and a Tourist Town

The population contributing to the sewershed was expected to fluctuate over the surveillance period due to tourism, weekday commuters, temporary workers, and quarantine etc. To monitor the population fluctuation, wastewater samples were collected from the WWTPs of City of Columbia (college town) and a tourist town over 10 time points (Table S2). The PARA load at each given time was calculated using PARA concentration and the daily flow volume reported in the metadata as in Eq. (3). The population at each given time was further estimated using the linear regression model built from Eq. (4) and the calculated PARA loads.

### 2.8 Relationships between SARS-CoV-2 load in wastewater and clinical prevalence

The weekly average of SARS-CoV-2 clinical case numbers in City of Columbia was collected from May to September 2021. *C*_*0*_ was calculated using metadata in Eq. (5); *C*_*2(PARA)*_ was calculated using the concentration of PARA in Eq. (10). SARS-CoV-2 concentration was normalized by *C*_*0*_ and *C*_*2(PARA)*_ depending on the scenarios: (1) SARS-CoV-2 load per capita normalized by metadata versus clinical cases normalized by metadata, (2) SARS-CoV-2 load per capita normalized by *C*_*2(PARA)*_ versus clinical cases normalized by metadata and (3) SARS-CoV-2 load per capita normalized by *C*_*2(PARA)*_ versus clinical cases normalized by PARA-estimated population using Eq. (12). Spearman’s correlation analysis was performed to examine the correlation between normalized SARS-CoV-2 concentration and one-week average clinical case numbers.

## 3. RESULTS

### 3.1 Relationships between biomarkers and population

Twenty-four samples collected from 12 WWTPs in the state of Missouri (Table 1) were used to explore the correlation between biomarkers and population using Eq. (2) or biomarker and population concentrations using Eq. (4). The linear regression models were fitted by either the biomarker concentration and population concentration ([*P*]) in Eq. (2) or biomarker loads and population in Eq. (4). The R square (R^2^) represents the variation of population/population concentration explained by the model. The Pearson’s correlation coefficient (*r*) represents the strength of the correlation.

The concentrations of CAF showed the highest correlation (Pearson coefficients, *r* = 0.810) with the population concentration in wastewater ([*P*]), followed by the concentrations of PARA (*r* = 0.774), pMMoV (*r* = 0.598), 5-HIAA (*r* =0.59), and CRE (*r* = 0.06) (Fig. 2 and Table S3). Log-transformation has been widely used to process the skewed data. It helps to decrease the variability of data and make data conform more closely to the normal distribution [37]. After log-transformation, the correlation coefficients were increased to 0.886 for CAF, 0.861 for PARA, 0.720 for 5-HIAA, and 0.707 for pMMoV (Fig. 3), however, it was not improved for CRE.

**Figure 2.**
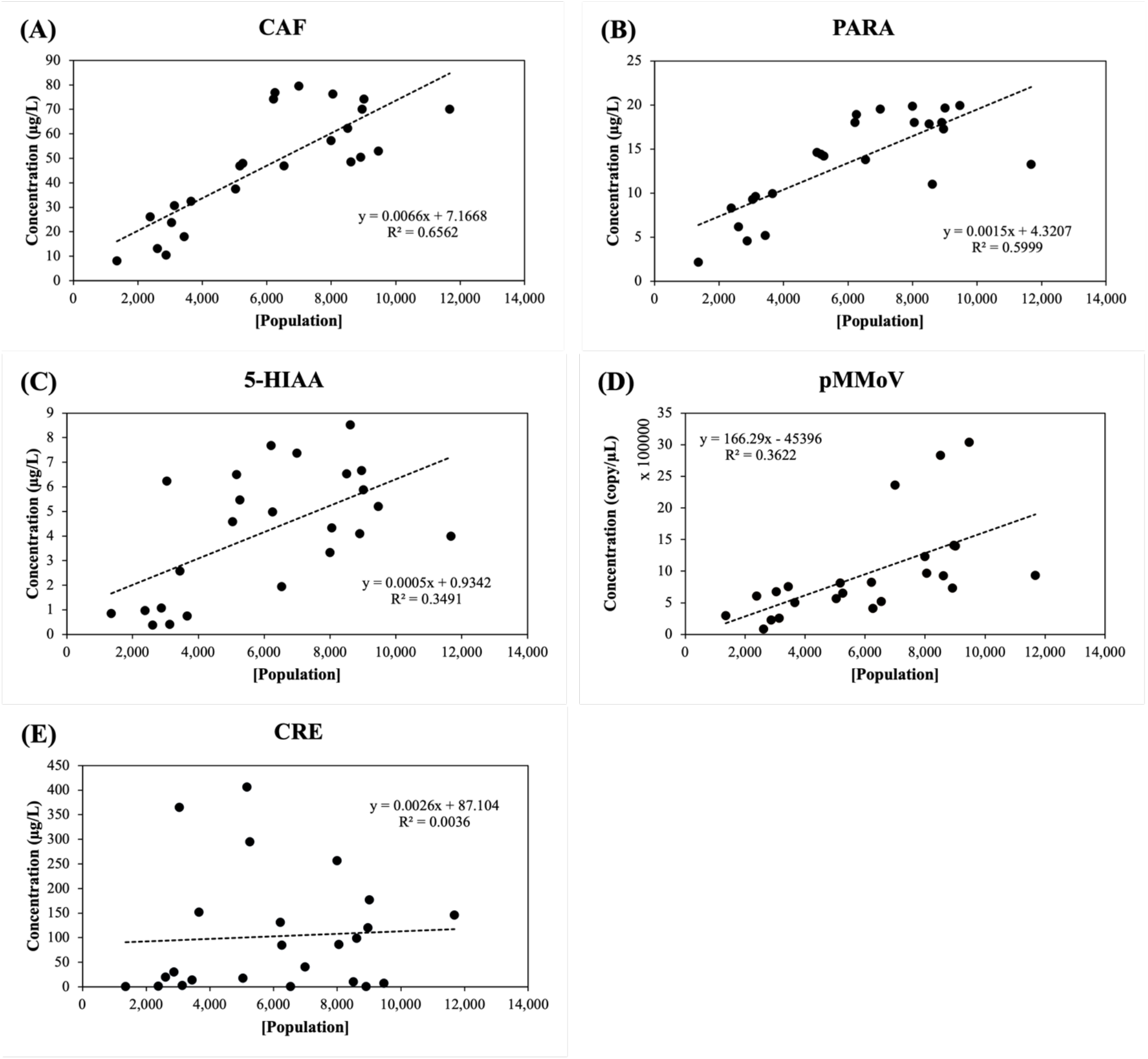
Population concentration [Population] versus biomarker concentration (mg/L) in the wastewater. (A) CAF: caffeine, (B) PARA: paraxanthine, (C) 5-HIAA: 5-hydroxyindoleacetic acid, (D) pMMoV: Pepper Mild Mottle Virus (E) CRE: creatinine. The concentrations of caffeine, paraxanthine, 5-hydroxyindoleacetic acid, and creatinine in 24 wastewater samples (Table 1) were determined by LC-MS/MS analysis and the Pepper Mild Mottle Virus concentration was determined by RT-qPCR as described in Methods and Materials. The population concentrations were calculated using the daily flow volume and population size in Eq. (1). The trendline (dashed line) was calculated using linear regression; R^2^ represented the percentage of the population concentration variation that is explained by the linear model.

**Figure 3.**
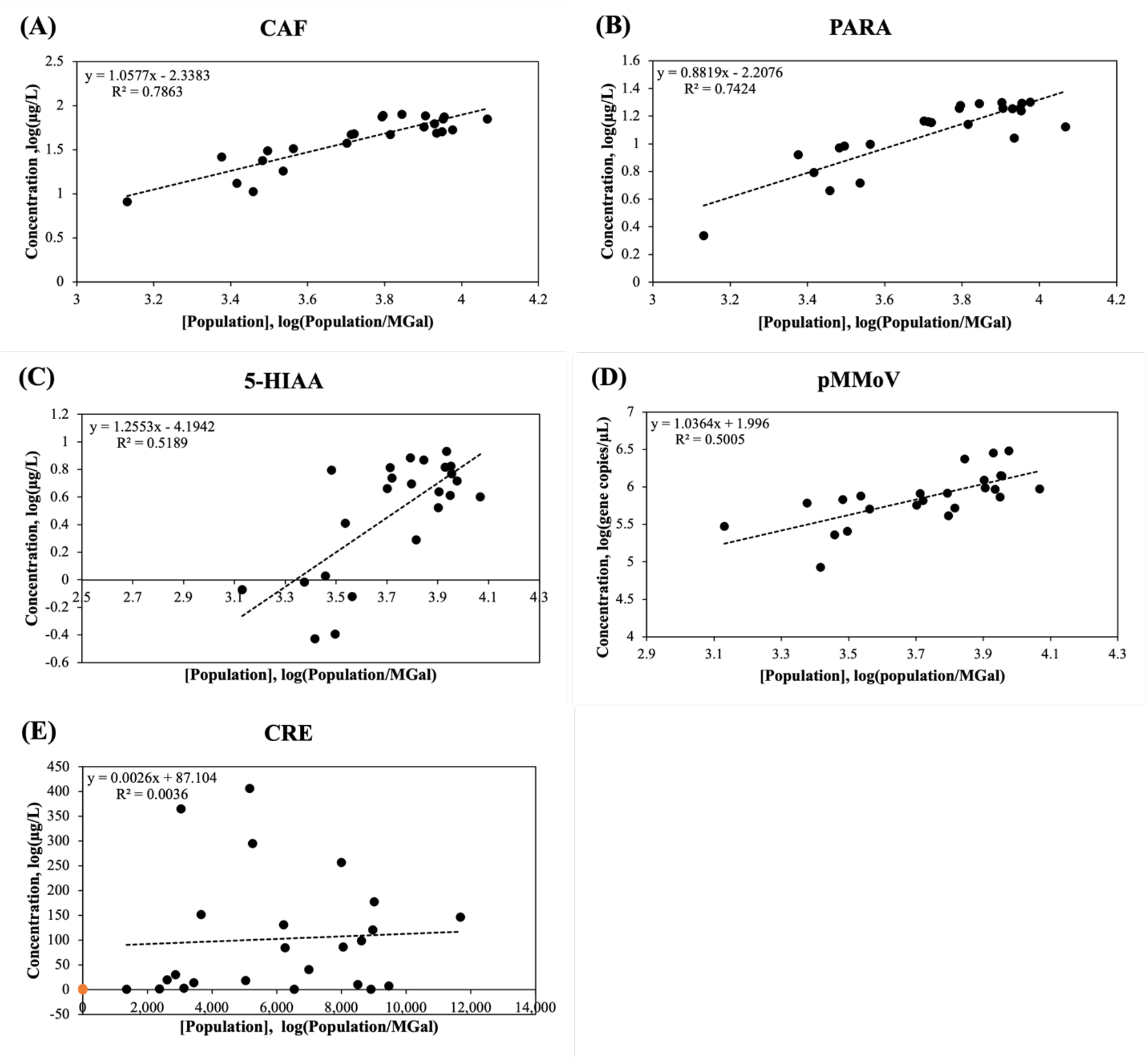
Log-transformed population concentration [Population] versus biomarker concentration (mg/L) in the wastewater. (A) CAF: caffeine, (B) PARA: paraxanthine, (C) 5-HIAA: 5-hydroxyindoleacetic acid, (D) pMMoV: Pepper Mild Mottle Virus (E) CRE: creatinine. The concentrations of caffeine, paraxanthine, 5-hydroxyindoleacetic acid, and creatinine in 24 wastewater samples (Table 1) were determined by LC-MS/MS analysis and the Pepper Mild Mottle Virus concentration was determined by RT-qPCR as described in Methods and Materials. The population concentrations were calculated using the daily flow volume and population size in Eq. (1). The trendline (dashed line) was calculated using linear regression; R^2^ represented the percentage of the population concentration variation that is explained by the linear model.

The daily load of CAF exhibited the highest correlation (*r* = 0.99) with population, followed by the daily load of 5-HIAA (*r* = 0.98), pMMoV (*r* = 0.98), PARA (*r* = 0.97), and CRE (*r* = 0.22) (Fig. 4 and Table S4). Similarly, log-transformation significantly improved the correlation of all five coefficients. The PARA and CAF daily load showed the highest correlation (*r* = 0.97 and 0.97, respectively) with the population, followed pMMoV load (*r* = 0.92), 5-HIAA load (*r* = 0.87), and CRE load (*r* = 0.33) after log-transformation (Fig. 5).

**Figure 4.**
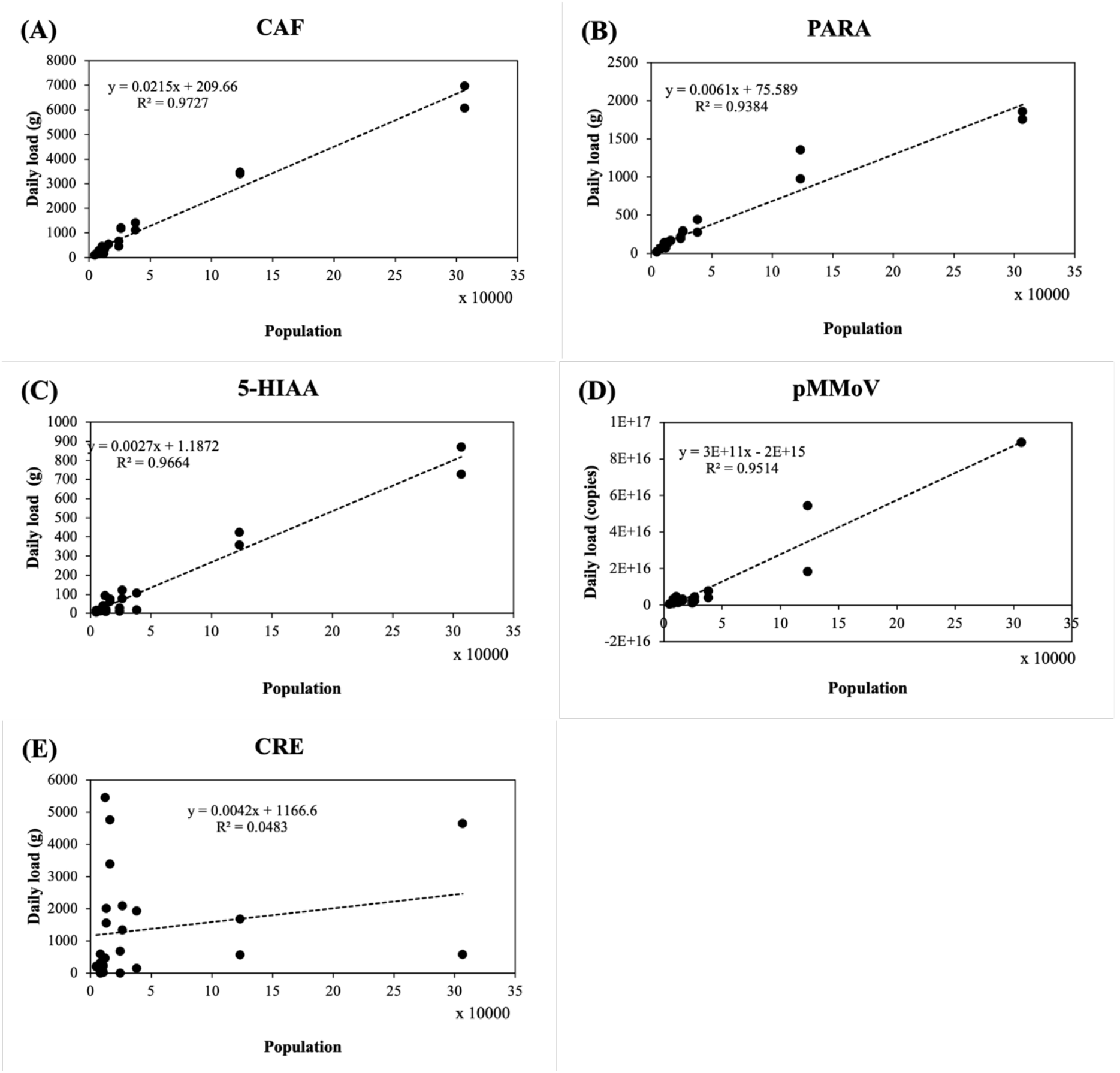
Population versus biomarker load in the wastewater. (A) CAF: caffeine, (B) PARA: paraxanthine, (C) 5-HIAA: 5-hydroxyindoleacetic acid, (D) pMMoV: Pepper Mild Mottle Virus (E) CRE: creatinine. The concentrations of caffeine, paraxanthine, 5-hydroxyindoleacetic acid, and creatinine in 24 wastewater samples (Table 1) were determined by LC-MS/MS analysis and the Pepper Mild Mottle Virus concentration was determined by RT-qPCR as described in Methods and Materials. The biomarker loads were calculated using the daily flow volume (million gallon, MGal) and biomarker concentrations in Eq. (3). The trendline (dashed line) was calculated using linear regression; R^2^ represented the percentage of the population concentration variation that is explained by the linear model.

**Figure 5.**
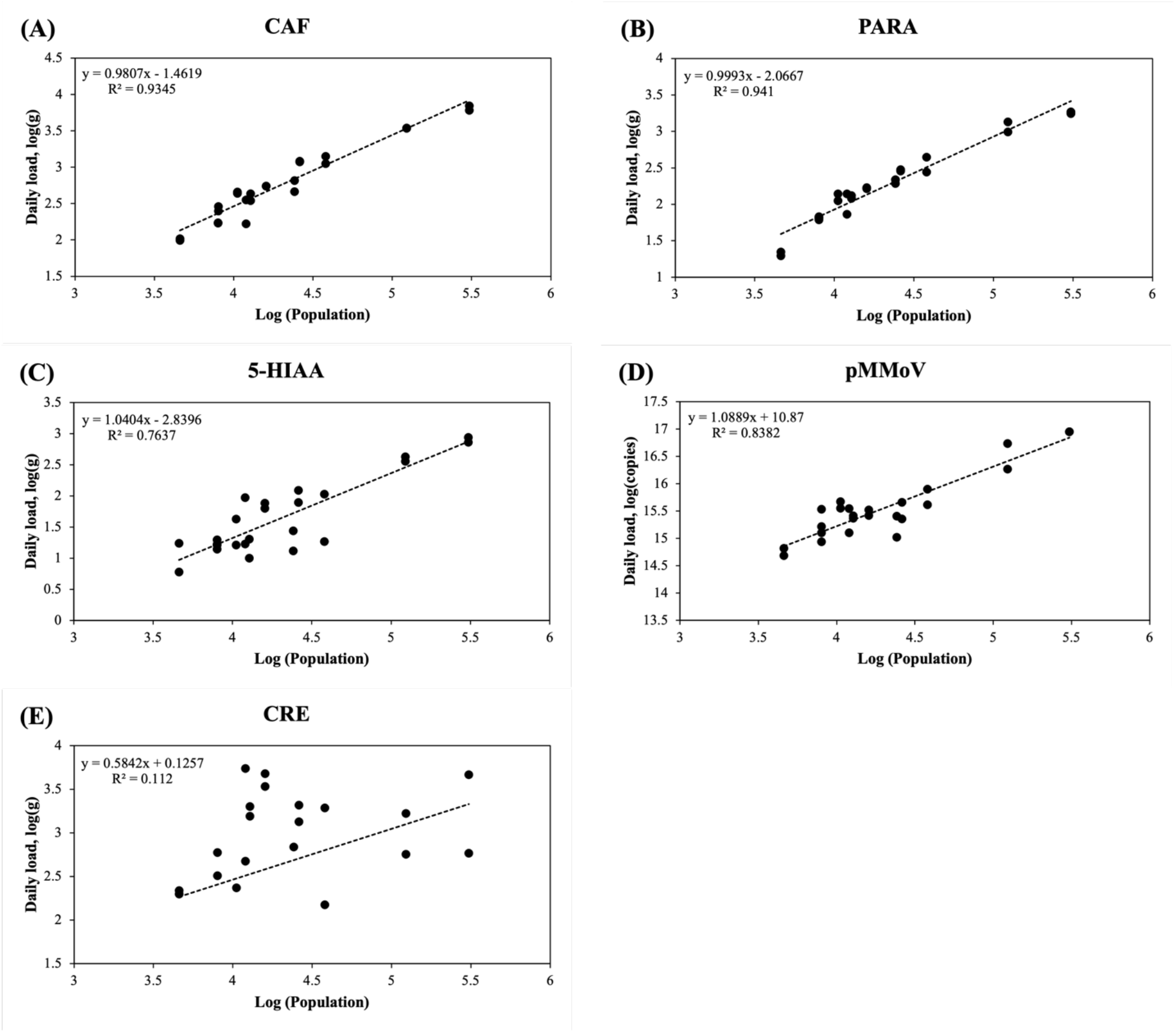
Log-transformed population versus biomarker load in the wastewater. (A) CAF: caffeine, (B) PARA: paraxanthine, (C) 5-HIAA: 5-hydroxyindoleacetic acid, (D) pMMoV: Pepper Mild Mottle Virus (E) CRE: creatinine. The concentrations of caffeine, paraxanthine, 5-hydroxyindoleacetic acid, and creatinine in 24 wastewater samples (Table 1) were determined by LC-MS/MS analysis and the Pepper Mild Mottle Virus concentration was determined by RT-qPCR. The biomarker loads were calculated using the daily flow volume and biomarker concentrations in Eq. (3). The trendline (dashed line) of each graph was generated using linear regression; R^2^ represented the percentage of the population concentration variation that is explained by the linear model.

### 3.2 Comparison of Normalization coefficients among Different Biomarkers

The normalization coefficient (*C*_*1(i)*_ or *C*_*2(i)*_) calculated from biomarker concentration were utilized to normalize SARS-CoV-2 viral load. A reliable biomarker for population normalization should achieve high precision and low variability, meaning that the normalization coefficient (*C*_*1(i)*_ or *C*_*2(i)*_ for biomarker *i*) should be comparable to *C*_*0*_ calculated from the population and daily flow volume derived from metadata. Hence, when the normalization coefficients from different biomarkers were standardized by *C*_*0*_ as fold change (*C*_*1(i)*_/*C*_*0*_), the closer to 1 (y=1) the fold change is, the higher precision and lower variability the biomarker obtains.

In the direct normalization approach, *C*_*1(i)*_ were calculated using the Eq (7) and biomarker concentrations. CAF outperformed other biomarkers resulting from the lower variation, and higher precision in comparison of the *C*_*1(i)*_ of all other biomarkers (Fig. 6 and Table S5). Most of *C*_*1(5-HIAA)*_ and *C*_*1(pMMoV)*_ among wastewater facilities showed variation above the baseline (y = 1), which could result in over-normalization of SARS-CoV-2. The relatively high variation of *C*_*1(5-HIAA)*_ and *C*_*1(pMMoV)*_ could over-normalize or under-normalize. The *C*_*1(CRE)*_ results were not included in this comparison due to its poor correlation with population. Therefore, the results suggested that the CAF should be the most suitable biomarker for the direct normalization approach, followed by PARA, 5-HIAA and then pMMoV at last.

**Figure 6.**
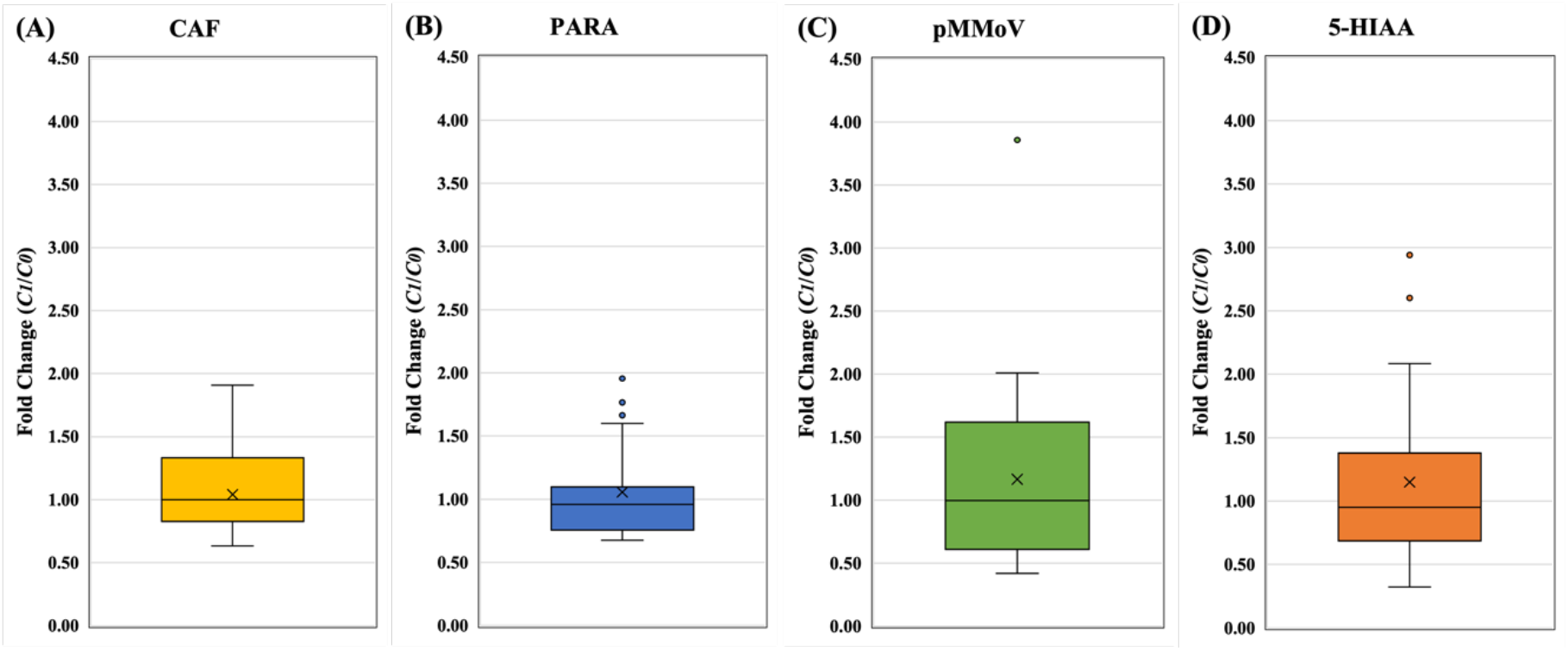
The fold changes of normalization coefficients from direct approach. A) CAF: caffeine, (B) PARA: paraxanthine, (C) pMMoV: Pepper Mild Mottle Virus, (D) 5-HIAA: 5-hydroxyindoleacetic acid. The normalization coefficients, *C*_*0*_ and *C*_*1(i)*_, of 24 wastewater samples (Table 1) were calculated using metadata and biomarker concentration in Eq. (5) and Eq. (7), respectively. The fold changes, *C*_*1(i)*_ divided by *C*_*0*_, were used to standardize *C*_*1(i)*_ for each biomarker at each WWTP. In the box plots, the upper whisker represents the maximum, the lower whisker the minimum; “X” represents the mean and open circles are the outliers. The data of CRE is not shown due to poor correlation between biomarker concentration and population concentration in wastewater.

In the indirect normalization approach, the normalization coefficients (*C*_*2(i)*_) were calculated with the data-transformed biomarker concentrations in Eq (9), followed by standardization by *C*_*0*_ and expressed as fold change (*C*_*2(i)*_/*C*_*0*_). The fold change (*C*_*2(PARA)*_/*C*_*0*_) of PARA outperformed other biomarkers due to its lower variation, and higher precision (Fig. 7 and Table S6). Among all biomarkers, CRE exhibited the highest variation and lowest precision. Thus, the most suitable biomarker for the indirect normalization approach would be PARA, followed by CAF, pMMoV and 5-HIAA.

**Figure 7.**
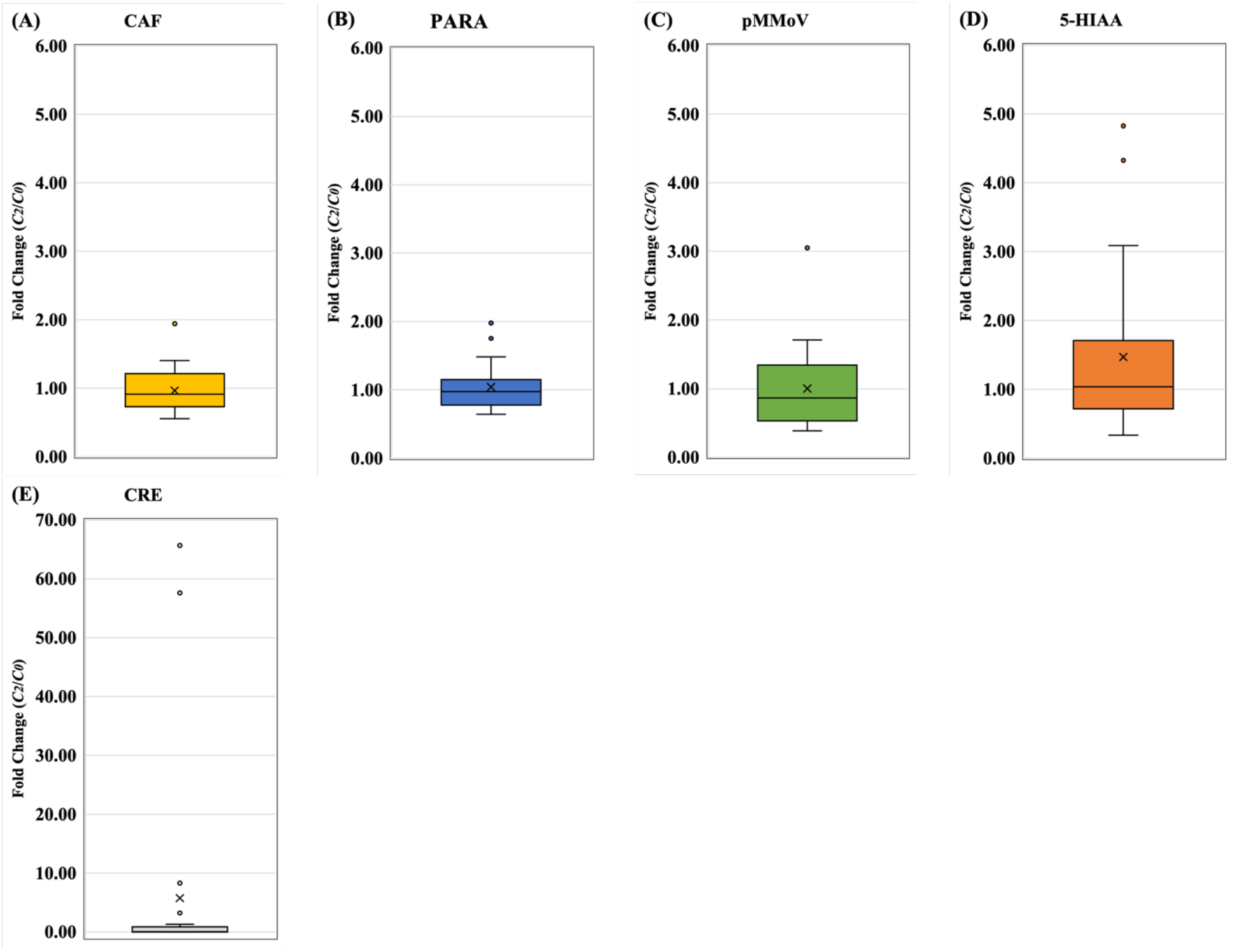
The fold changes of normalization coefficients from indirect approach. A) CAF: caffeine, (B) PARA: paraxanthine, (C) 5-HIAA: 5-hydroxyindoleacetic acid, (D) pMMoV: Pepper Mild Mottle Virus (E) CRE: creatinine. The normalization coefficients, *C*_*0*_ and *C*_*2(i)*_, of 24 wastewater samples (Table 1) were calculated using metadata and biomarker concentration in Eq. (5) and Eq. (10), respectively. The fold changes, *C*_*2(i)*_ divided by *C*_*0*_, were used to standardize *C*_*2(i)*_ for each biomarker at each WWTP. In the box plots, the upper whisker represents the maximum, the lower whisker the minimum; “X” represents the mean and open circles are the outliers.

### 3.3 Normalization of SARS-CoV-2 load per capita

The SARS-CoV-2 loads normalized by biomarkers (copies/person) were directly calculated by multiplying the viral concentrations with the normalization coefficient of the corresponding biomarker. Fig. 8 demonstrated the biomarker-normalized viral per capita of each selected facility in the State of Missouri for the week of January 19^th^ and week of January 23^rd^, 2021. Among all the facilities, the community within BROOK sewershed was identified as the most vulnerable community due to the highest viral loads per-capita (Fig. 8).

**Figure 8.**
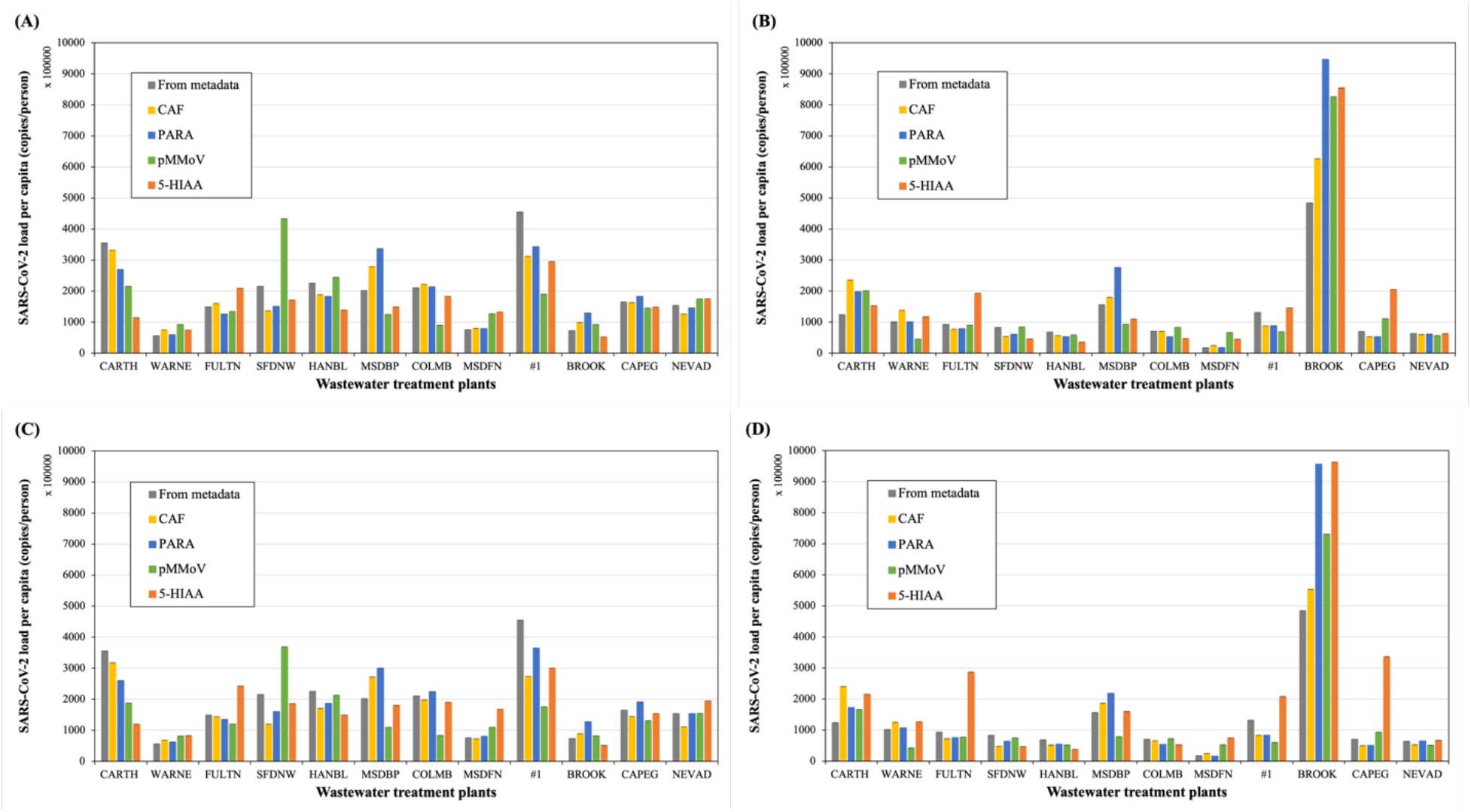
The normalized SARS-CoV-2 load per capita by biomarkers using either direct or indirect approaches at WWTPs. The direct normalization approach was applied to 12 samples collected in the week of (A) January 19^th^ and (B) January 23^rd^. The indirect approach was applied to 12 samples collected in the week of (C) January 19^th^ and (D) January 23^rd^. (Grey: Metadata, yellow: CAF, blue: PARA, green: pMMoV, orange: 5-HIAA; error bars showed standard deviation, n=4). The SARS-CoV-2 load per capita was normalized using the average of duplicated N1 and N2 concentrations at each WWTP and the normalization coefficients of each biomarker in Eq. (7) for direction approach in (A) and (B), or in Eq. (10) for indirect approach in (C) and (D). The viral loads were normalized using metadata in Eq. (5) and included in all graphs for comparison. The data of CRE was not shown due to its poor correlation with population.

### 3.4 Validation of normalization coefficients

Based on the value of fold change, CAF and PARA achieved the lowest variability and highest accuracy, and precision (Figures 6 and 7). These normalization approaches were further validated the using wastewater samples collected from 64 WWTPs in the State of Missouri in May 2021 (Table S1). The normalization coefficients, *C*_*1(CAF)*_, *C*_*1(PARA)*_, *C*_*2(CAF)*_ *and C*_*2(PARA)*_, for each WWTP was calculated using the established regression functions between CAF/PARA and population (Table S3 and S4) without metadata. These coefficients were normalized by *C*_*0*_ derived from metadata to assess the fitness, precision, and variability.

There was no significant difference between the normalization coefficients of CAF and PARA when the direct approach or indirect approach was applied (Fig. 9). The fold changes of CAF and PARA from direct and indirect approach were close to 1 (high precision and low variability). These results not only consistent with the results shown in Figure 4 and 5 but also indicated that the regression functions developed in this study could be used for normalizing SARS-CoV-2 load without metadata in the future.

**Figure 9.**
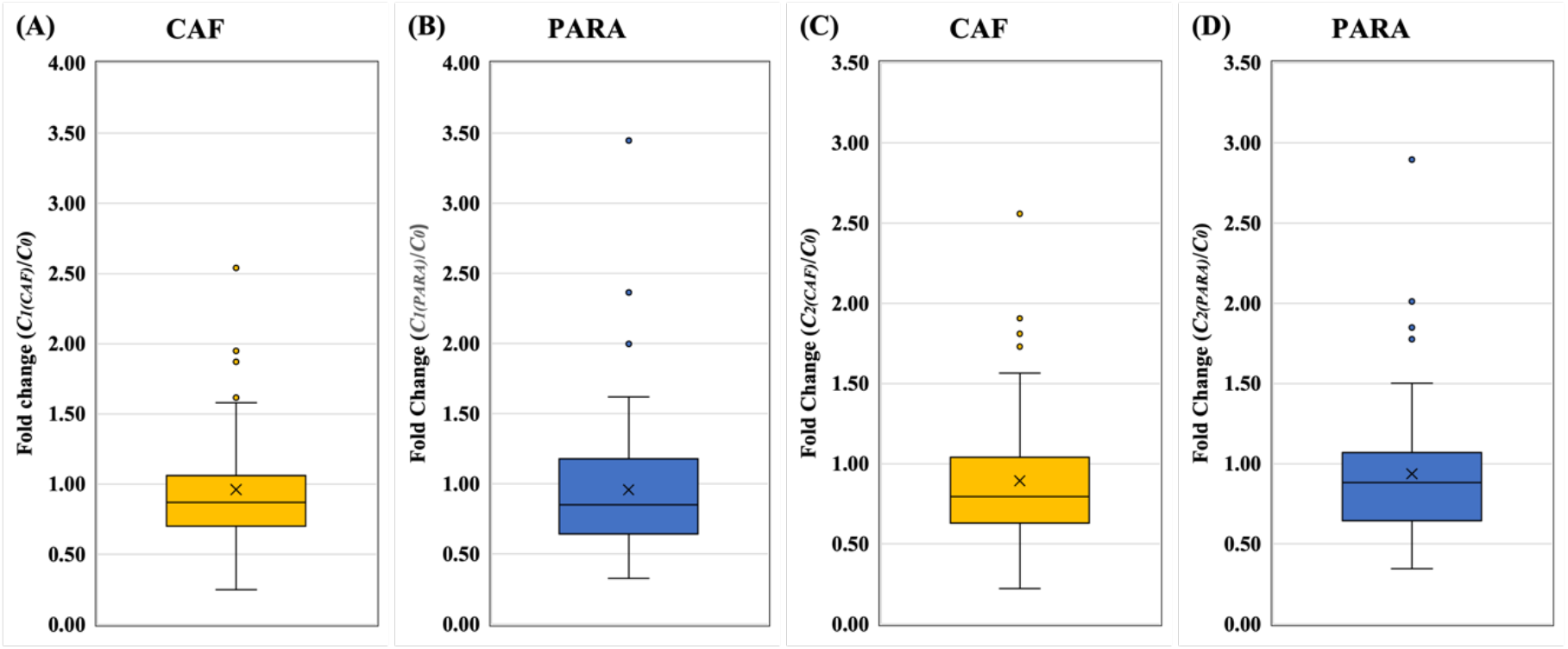
Validation of normalization approaches. The direct approach for (A) CAF and (B) PARA and the indirect approach for (C) CAF and (D) PARA were applied and shown for validation. In the box plots, the upper whisker represents the maximum, the lower whisker the minimum; “X” represents the mean and open circles are the outliers. The PARA and CAF concentrations in 64 wastewater samples collected from WWTPs in the State of Missouri (Table S1) were quantified by LC-MS/MS, and the normalization coefficients, *C*_*0*_, *C*_*1(i)*_ and *C*_*2(i)*_, were calculated as described in Methods and Materials. The fold changes (*C*_*1(i)*_ /*C*_*0*_ or *C*_*2(i)*_ /*C*_*0*_) were used to standardize *C*_*1(i)*_ and *C*_*2(i)*_.

### 3.5 Estimation of real-time population contributing to the wastewater

The precision of real-time biomarker-estimated populations were assessed by fitting regression models with the biomarker loads using R program. PARA achieved the highest adjusted R square, followed by CAF, 5-HIAA, pMMoV and CRE (Table 4). PARA showed the lowest mean square error (MSE), which is the parameter used for assessing the prediction accuracy by the developed model and it was increased in the order of CAF, pMMoV, 5-HIAA and CRE. Again, PARA obtained the lowest 5-fold cross-validation MSE, suggesting that PARA is the most suitable biomarker for estimating the population.

**Table 4.** Estimation of population using biomarker loads

| <sup>a</sup>Biomarkers | P value | Adjusted R<sup>2</sup> | <sup>a</sup>MSE | <sup>c</sup> *k*-fold Cross-validation MSE |
| --- | --- | --- | --- | --- |
| CAF | 0.00 | 0.938 | 0.0723 | 0.0251 |
| PARA | 0.00 | 0.9404 | 0.0516 | 0.0182 |
| 5-HIAA | 0.00 | 0.8351 | 0.6124 | 0.1065 |
| pMMoV | 0.00 | 0.9043 | 0.5125 | 0.0501 |
| CRE | 0.10 | 0.1189 | 0.9400 | 0.2517 |

<sup>a</sup> The biomarker loads, and population were transformed using log10.
<sup>b</sup> MSE: mean square error.
<sup>c</sup> *k*-fold Cross-validation was performed when *k*=5 and averaged MSE was calculated.

To accurately normalize SARS-CoV-2 loads per capita over time, the populations at a college town and a tourist town were estimated using the PARA concentrations in wastewater samples collected through May to early September in 2021. When the daily flow volume was available, the real-time population was predicted by the biomarker loads using the established biomarker loads vs. populations regression functions in Eq. (3) (Table S4). The results showed the real-time population dynamic of population at City of Columbia, especially in late May, August, and early September (Fig. 10A). The variation of estimated populations in Columbia were from - 36% to 8% compared to the population reported in Metadata. The change in the real-time population from May to early September in a tourist town were observed in similar pattern (Fig. 10B).

**Figure 10.**
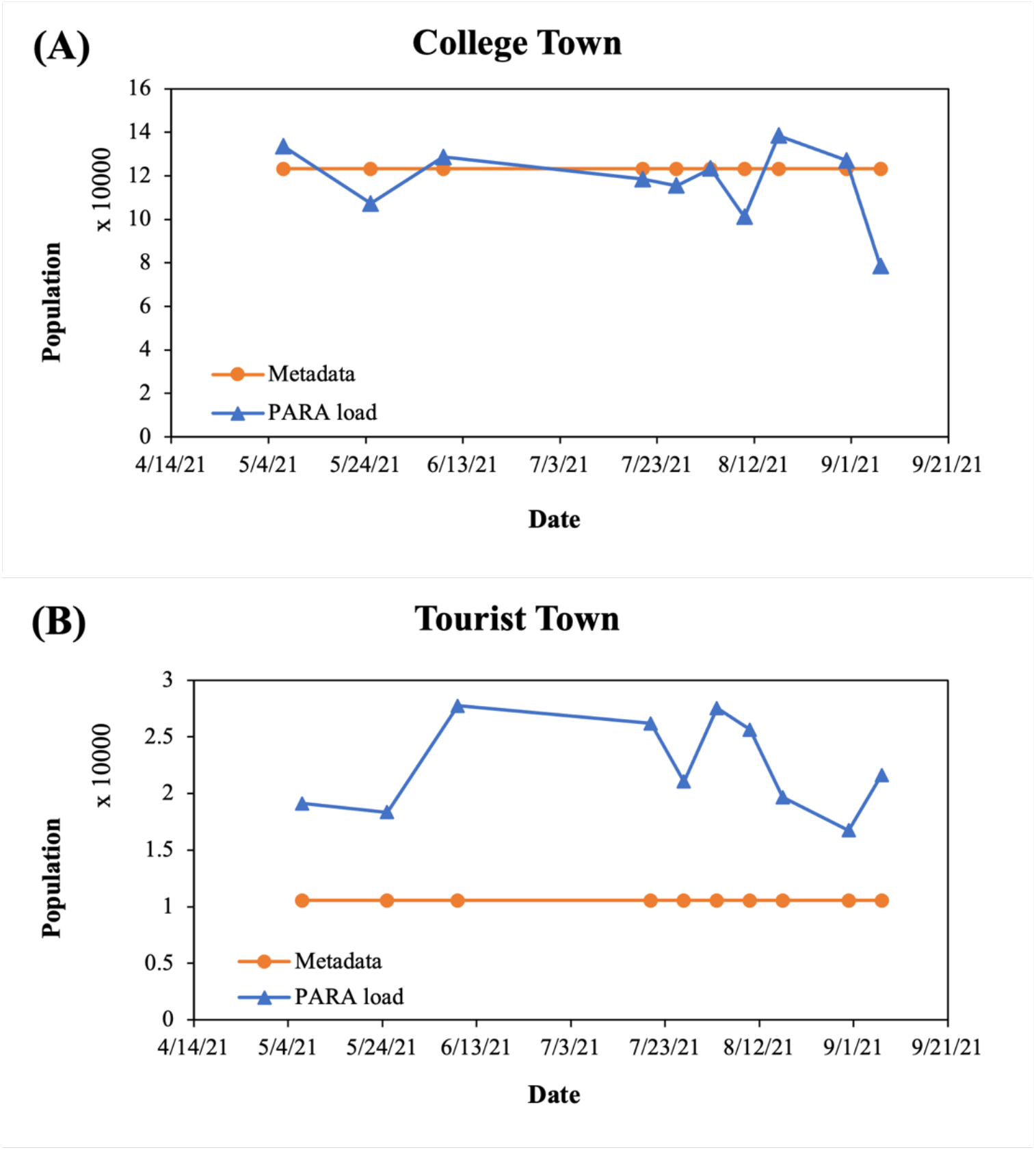
Estimation of real-time population in the college town and the tourist town. (A) College town (B) Tourist town (blue triangle: population estimated using PARA, orange circle: population reported by Metadata). The PARA concentrations in 10 wastewater samples collected from WWTPs in City of Columbia and a tourist town (Table S2) were quantified by LC-MS/MS as described in Methods and Materials and further converted to daily PARA load using daily flow volume from metadata. The population was estimated using the daily PARA load using the developed regression function (Table S4).

### 3.6 Correlation between SARS-CoV-2 load per capita and clinical prevalence

It was demonstrated in Fig. 11 that the relation between SARS-CoV-2 levels in the wastewater and clinical cases could be mispresented without a proper normalization using a reliable population marker. This is mainly attributed to that the population in the City of Columbia was constantly fluctuating over the surveillance period (Fig. 10A). The Spearman’s rank correlation was performed to understand the correlation between viral loads and prevalence data [38]. Spearman’s correlation coefficient, *rho*, represents strength of the correlation between viral loads and prevalence data.

**Figure 11.**
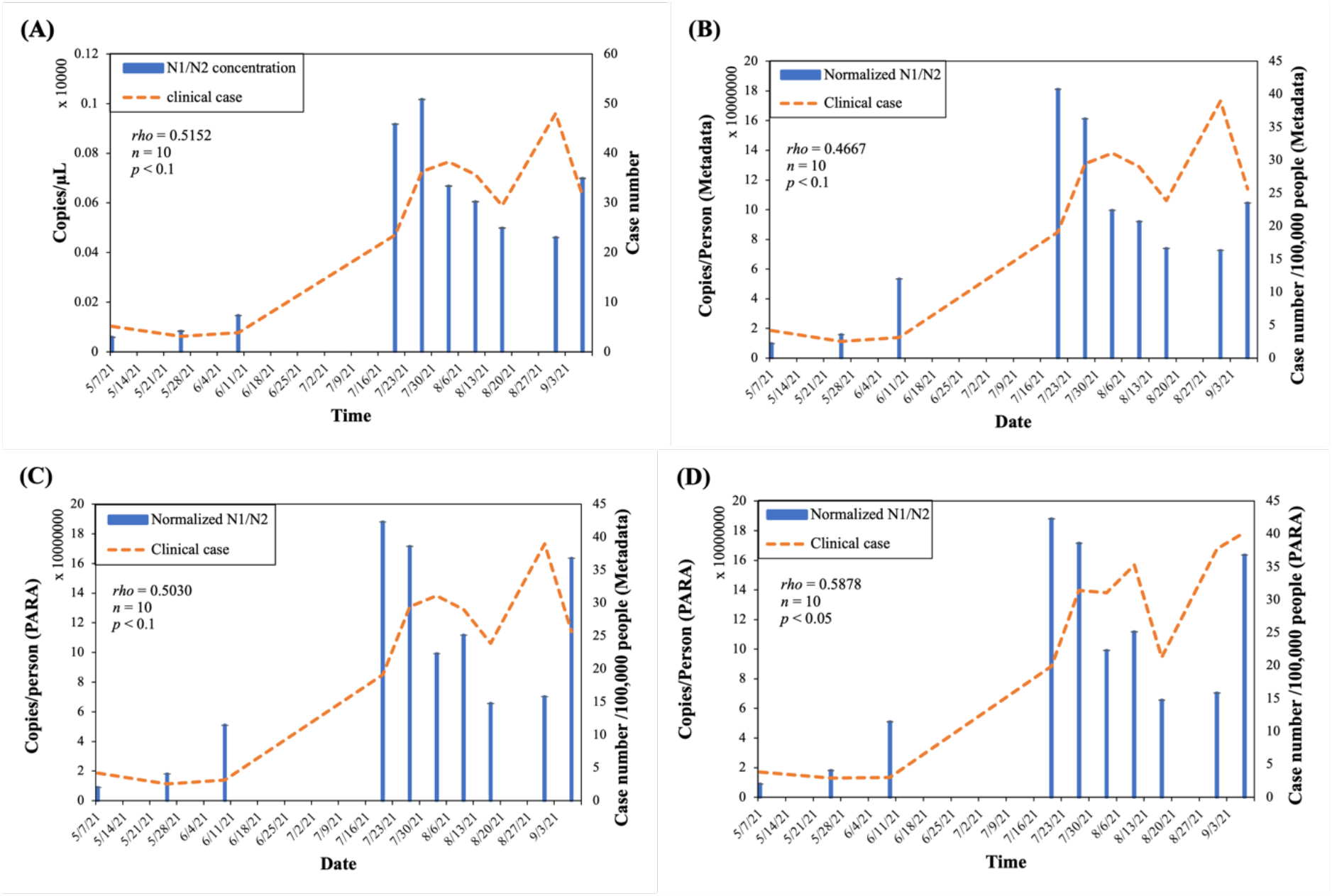
The correlation between normalized SARS-CoV-2 loads in wastewater and the clinical reported case numbers. (Orange dashed line: clinical case, blue solid bar: normalized N1/N2 average concentration/load). The PARA concentrations in 10 wastewater samples collected from WWTP in City of Columbia (Table S2) were quantified by LC-MS/MS as described in Methods and Materials and applied in Eq. (10) to normalize viral load using indirect approach. (A) Viral concentrations and clinical cases before normalization (B) Both viral load per capita and clinical cases normalized using metadata. (C) Viral load per capita normalized by PARA load and clinical cases normalized by Metadata (D) Both viral load per capita and clinical cases normalized by PARA loads. The Spearman’s correlation was performed to examine the correlation between normalized SARS-CoV-2 and clinical case numbers; *rho* represented the strength of the correlation.

For instance, the correlation between the average weekly case number and the SARS-CoV-2 concentration over time was insignificant (*rho* = 0.5152, *p* < 0.1) before normalization (Fig. 11A). The *rho* was reduced to 0.47 (*p* < 0.1) after the viral concentration and clinical case number were both normalized by the fixed population from the metadata (through population census) (Fig. 11B). Similarly, as the viral concentration normalized by PARA-estimated population plotted against the clinical case numbers normalized by metadata, *rho* dropped to 0.50 (*p* < 0.1) (Fig. 11C). In contrast, when both viral load and clinical case number were properly normalized using PARA, the correlation was positive and moderate (*rho* = 0.59, *p* < 0.05) (Fig. 11D).

## 4. DISCUSSION

### 4.1 Population Biomarker selection

Although the United States Centers for Disease Control (CDC) has recommended using pMMoV as population fecal biomarker to normalize SARS-CoV-2 concentrations, our findings suggested that the chemical marker, PARA, is more reliable population biomarker, due to its 1) better population indicators with higher accuracy, lower variability and higher temporal consistency, 2) very limited exogenous sources, 3) high extraction efficiency with low variability, 3) high stability, 4) resistant to chemicals in the wastewater, and 5) low sample volume requirement with simple sample preparation process.

The log-transformed PARA daily load demonstrated better correlation with population (*r* =0.97) as compared to pMMoV (*r* = 0.92, Fig.4). For both direct and indirect normalization approaches, PARA always outperform pMMoV and showed more accurate normalization coefficients with lower variability.

Pepper Mild Mottle virus (pMMoV), a single stranded RNA virus commonly found in the diet, has been an attractive marker used for human fecal normalization since it has high concentrations in sewage and can be used simultaneously quantified as the targets SARS-CoV-2 viral nucleic acid using the multiplex platforms. The PMMoV is constantly excreted by human and unaffected by seasonal variations in wastewater [3,19,39]. Our findings demonstrated that this genetic biomarker showing positive correlation with population (*r* =0.92, Fig. 4), which is consistent with the findings reported by D’Aoust et al. [40].

However, the exogenous sources [16,18], variation in the extraction rates [41], and relatively short half-life as compared to several chemical biomarkers have been the main drawbacks of pMMoV. These drawbacks might have contributed to its lower correlation coefficients as compared to CAF and PARA in this study. The pMMoV has been widely detected in the groundwater, irrigation water and surface water (rivers, ponds). For example, Rosiles-González et al. detected pMMoV in the groundwater during the raining season and the concentration of pMMoV didn’t correlate with other fecal indicator, such as *E. coli*. Asami et al. also reported similar results that pMMoV concentrations changed between dry and wet seasons in dirking water sources, whereas *E. coli* counts remained unchanged [42]. The pMMoV was also detected in 100% of river water samples collected near North Rhine Westphalia region (NRW), one of the most populated areas in Germany, at concentrations ranging from 10^3^–10^6^ genome copies GC/L, while the concentrations of pMMoV in wastewaters is often ranging from 10^6^ to 10^10^ GC/L [43]. Previous studies also reported the presence of pMMoV in pond and irrigation waters. Kuroda et al. reported that pMMoV was detected in 91% of samples collected form the pond waters, with concentrations ranging from non-detectable to 1.2 × 10^5^ GC/L. Similarly, pMMoV was found in 100% samples collected from the irrigation waters [44]. In addition, recently, several SARS-CoV-2 wastewater surveillance projects in the U.S. have reported the increased levels of pMMoV after the major stormwater events. Further investigation suggested the potential exogenous sources of the pMMoV from agricultural soils, suspended sediments and fertilizers (personal communication).

Variations in the extraction rates of pMMoV that have been widely reported is another drawback [45–47]. Feng et al. reported a recovery of 45±26% pMMoV using direct extraction with HA filters. The pMMoV was also poorly correlated with the recovery of the SARS-CoV-2 enveloped virus [40]. Similarly, Kato et al. reported a wide variability of the pMMoV recovery efficiencies with typical recovery rates only greater than >10% when concentrating using electronegative filters [47]. The high variability among different concentration techniques for pMMoV analysis, including direct extraction, HA filtration, filtration with bead bearing, PEG precipitation, and ultrafiltration have been illustrated by LaTurner et al.[46]. The coefficient of variation (%CV) for these concentration techniques range from 25.9% to 49.8%. Feng et al. reported that the variability in the pMMoV extraction rates might have contributed to the decreased correlation coefficient between the normalized SARS-CoV-2 concentration and the clinic cases in most of WWTP facilities reported by previous studies [45]. Among the genic fecal markers, although pMMOV has demonstrated a less variable RNA signal compared to Bacteroides 16S rRNA or human eukaryotic 18S rRNA, the variability of pMMOV assay could be significant with Ct variance from 1.18 to 1.34 [40,45].

Although pMMoV has been known to be persistent in the soils, the results of an incubation study suggested that the half-lives of the pMMoV in river water ranges from 7 to 10 days, depending on the temperatures. At 0°C, PMMoV showed 1.1 log10 reduction (7.9 % remaining) after 21 days of incubation in river water with PMMoV half-life of about 7 days. At 25C, PMMoV showed 3.7 log10 reduction (0.02 % remaining) after 21 days of incubation in river water with a half-life of about 10 days. As compared to more stable CAF and PARA, the relative short half-life of the pMMoV suggest that the pMMoV assays need to be completed within 1 week after the samples are received, even they are properly stored at 4C. Moreover, despite that no inhibition observed in the one step RT-qPCR assay in our study, RT-qPCR inhibition have been reported by several studies [47]. Quality control internal standards, and dilution protocols are often required to account for any PCR inhibition. Incorporation of the internal positive control, such as a modified targeted gene sequence or CGMMV are often required to correct the variation in the extraction efficiency plus any potential inhibition [47].

On the other hand, both CAF and PARA, the major metabolite of caffeine, exhibited good, consistent high recovery rates and high stability in the wastewater as compared to pMMPoV (Table 5). The average recovery rates of CAF and PARA in our study were 101% and 92% with standard deviation of ±7% and ±3%, respectively, similar to 73% to 109% for CAF and its metabolites reported by Driver et al. [24]. Both CAF and PARA were found to be relatively stable in the sewer system [48]. The CAF and PARA have several unique characteristics that are critical to serve as the reliable chemical fecal population markers. They are highly soluble in water (13 g L^−1^) with a very low hydrophobicity (octanol-water coefficient log K_ow_ = −0.07), insignificant volatility and its half-life is about 10 years [49–52]. Due to the high polarity and water solubility, CAF and PARA will less likely to adhere to the solids fraction of wastewaters via electrostatic and/or hydrophobic partitioning effects as the pMMoV biomarker described by Armanious et al.[53]. As the wastewater stored at -20°C, the PARA could be stable for at least 4 weeks or more [25,48]. With the new modified direct methanol dilution extraction protocol (50% methanol), we anticipate that the CAF and PARA extracts could be stable beyond several months when they are stored at -20 C° under the 50% methanol sterilized solution [54].

**Table 5.** Comparison of selected biomarkers in this study.

|  | CAF | PARA | 5-HIAA | pMMoV | CRE |
| --- | --- | --- | --- | --- | --- |
| **Stability in Wastewater** | Stable [20,48] | Stable [48] | Stable [58] | Poor | Poor [57] |
| **Storage stability** | Stable<br>> 40 days | Stable<br>> 40 days | - | Poor<br>(half-life 6-10 days) | - |
| **Recovery/**<br>**Extraction Rate** | <sup>a</sup>101±7% | <sup>a</sup>92±3% | <sup>a</sup>78 ± 19% | 10%-45%± 40%-50% | <sup>a</sup>123±31% |
| **LOD** | <sup>b</sup>1.06 µg/L | <sup>b</sup>0.72 µg/L | <sup>b</sup>14.74 µg/L | 100 copies/µL | <sup>b</sup>1.19 µg/L |
| **Signal inhibition** | No | No | No | Sensitive | No |
| **Concentration in wastewater** | 47.3 ± 22.9<br>µg/L | 4.2 ± 2.5<br>µg/L | 13.5 ± 5.5<br>µg/L | 959920 ± 773834<br>copies/µL | 102.8 ± 120.4<br>µg/L |
| **Sample Volume** | 1.5-2 mL | 1.5-2 mL | 25-50 mL | 50 mL | 1.5-2 mL |
| **Sample Preparation time**<br>**(for 12 samples)** | 30 mins | 30 mins | 2-3 hours | 3-4 hours | 30 mins |
| **Analysis time** | 15 minutes per sample | 15 minutes per sample | 15 minutes per sample | 2 hours for 64 samples | 15 minutes per<br>sample |
| **Other exogenous sources** | Disposal of the coffee<br>or caffeinated products | Microbial degradation<br>of caffeine (small<br>amount)[56] | - | Ground water,<br>agriculture soils,<br>fertilizers. | - |

<sup>a</sup> The recovery rate was calculated from the isotope fortified in wastewater samples.
<sup>b</sup> The limit of detection of LC-MS/MS method as described in the Material and Method.
<sup>c</sup> The limit of detection of RT-qPCR assay as described in the Material and Method.

In addition, the sample volume required for analysis for PARA is less than 2 mL (0. 1mL with a modified methanol extraction protocol), that is significantly less than 25-50 mL sample volume required for pMMoV analysis (Table 5). Another advantage for using PARA as the fecal marker is that it required less sample preparation time and processes. An average sample preparation time for PARA analysis was less than 30 minutes/6 samples, with new modified methanol extraction protocols, it could be further reduced to 10 minutes/6 samples, while the sample preparation time (e.g., extraction and concentration) for pMMoV analysis often takes approximate 3 hours. Most importantly, unlike CAF and pMMoV, PARA is the metabolite product generated through the human consumption of the caffeinated products (coffee, tea and caffeinated drinks), indicating that human is the major source contributing PARA in the wastewater. In humans, 80% of caffeine is metabolized into paraxanthine [55]. The production of the PARA could be also attributed to the microbial degradation of caffeine in the environments, however since it is not the predominant microbial degradation pathway, the amount of PARA produced through this process is very limited [56]. Therefore, we could assume that PARA loading in WWTP was mostly generated through human consumption of caffeine. Unlike the PARA, the CAF loading might result from discarded caffeinated products, and therefore, make CAF less desirable population biomarker.

Other biomarkers do not meet the criteria of population biomarker. Creatinine, the metabolite of muscle, didn’t correlate with population, consistent with the results reported by Thai et al. [57,58]. The poor correlation could be due to its instability in wastewater treatment designs and processes, high variance of intra- and extra-individual excretion [57,59]. The 5-HIAA, one of the major metabolites of serotonin, correlated with population well and it has been reported to be stable in wastewater [58]. Nevertheless, the low concentrations in the wastewater and the observed coeluted interferences in the LCMSMS analysis, the time required for sample preparation and cleanup, particular the time-consuming concentration and cleanup processes through solid-phase extraction (SPE), make the 5-HIAA not an ideal marker candidate for real-time and rapid analysis. In addition, a sensitive tandem mass spectrometer is the only option for quantifying the 5-HIAA in the wastewater due to its low sub-ppb to ppb concentration range, while CAF and PARA could be quantified by other less-expensive alternative analytical techniques, such as gas chromatography–mass spectrometer (GC-MS), high-performance liquid chromatography coupled with photodiode-array detector (HPLC-PDA) due to their much higher concentrations in the wastewater sample[60,61].

### 4.2 Normalization of SARS-CoV-2 load and method validation

The utility of chemical biomarkers for human fecal normalization in SRAS-CoV-2 WBE surveillance was so far very limited. This study investigated several alternative chemical population biomarkers in SARS-CoV-2 WBE. These chemical population biomarkers were extracted and analyzed by LCMSMS. The concentrations of biomarkers were applied to the exercise in correlation with population to generate their normalization coefficient. The SARS-CoV-2 loads per capita were normalized using the normalization coefficient of each chemical population biomarker. Both direct and indirect approaches aimed at precisely estimating the population concentration (population per MGal) that would be applied in the following determination of the viral load per capita (Fig. 3 and 5). The normalization coefficient calculated from different biomarker can be compared and evaluated before SARS-CoV-2 concentration involved. Most importantly, our normalization approaches can be proceeded without daily flow volume and the size of the population using the regression functions established in this study (Table S3 and S4). However, the traditional normalization requires the information of the daily flow volume and population size. The SARS-CoV-2 concentration was converted to mass using daily flow volume, followed by being divided by population served by the WWTP (Fig. 1A) to obtain viral loads per capita.

In our normalization approaches, the parameter fold changes, the normalization coefficients (*C*_*1*_ and *C*_*2*_) standardized by *C*_*0*_ (from metadata), were utilized to evaluate the fitness of the biomarkers for each normalization approach as compared to the traditional method. The fold change that is closes to 1 indicates the highest accuracy. For example, in the direct approach, fold changes for CAF and PARA were 1.041±0.3111 (mean±standard deviation) and 1.057±0.389, respectively, and 0.967±0.324 and 1.042±0.341, respectively, in the indirectly approach.Both CAF and PARA showed high accuracy and low variability in either approach. On the contrary, the fold changes of 5-HIAA and pMMoV showed significantly difference by between two approaches. The 5-HIAA fold change was 1.150±0.661with the direct approach but 1.470±1.144 in the indirect approach, whereas pMMoV performed better (1.003±0.586) with the indirect approach than (1.166±0.737) in the direct approach. (Table S5 and S6). The high accuracy and low variability by CAF and PARA are possibly attributed to high reproducibility of the analysis, high recovery rates, stability of these molecules, and low adsorption affinity to the solids fraction of wastewaters.

Furthermore, the regression functions established by CAF and PARA in our two approaches can be utilized to determine the population concentration in the long-term monitoring without knowing daily flow volume and population size in the future WBE applications. The normalization approaches were validated using additional 64 samples collected from May 2021 (Table S1) with the established regression functions of CAF and PARA. The fold changes of CAF and PARA from these additional 64 samples obtained high precision and low variation in both direct and indirect approaches (Fig 9), consistent with our results from the developed models (Fig 6 and 7).

This is the first study to normalize the SARS-CoV-2 load with biomarker estimated population and to accomplish viral load per capita with a universal unit ¾ copies/person. Most of the previous studies utilized biomarker to normalize SARS-CoV-2 concentrations but got a unitless results (eg. N1/N2 copies/copies of genetic biomarker). Green *et al*. reported the ratio of SARS-CoV-2:crAssphage in the wastewater; N1 or N2 copies/copies of biomarker (pMMoV, BCoV, HF183, crAssphage, and Bacteroides rRNA) in the wastewater were reported by Feng *et al*.; Greenwald *et al*., and Ai *et al*.; D’Aoust *et al*. and Wolfe *et al*. presented copies/copy of pMMoV in solids (Table S9). Nevertheless, the biomarker-estimated population should be incorporated into surveillance programs, so the normalization can reflect the real viral per capita to be compared over time and cross facilities and be further utilized for predicting the trend of COVID-19 prevalence.

### 4.3 Relationship among estimated real-time population, SARS-CoV-2 in wastewater and prevalence

The fluctuations in the population posed a challenge to WBE long-term monitoring [3]. If the population contributing to the sewershed is expected to constantly change over the surveillance period (due to tourism, weekday commuters, temporary workers, etc.), population normalization is extremely critical to interpret SARS-CoV-2 concentrations and predict the trend and the infected population over time. We successfully demonstrated the utility of PARA for gauging small-area populations in real-time and captured population dynamics in a college town and a tourist town (Fig. 10) resulting from PARA gave the highest adjusted R square with lowest MSE and 5-fold cross validation MSE in the population predicting model (Table 4). Our findings directly corresponded the fluctuations in the population due to seasonal activities in these tourist town and university community, such as the summer breaks, holidays (e.g., Labor Day weekend in September) and tourisms.

We strongly believe that population dynamic should be taken into consideration when the clinical cases are normalized for long-term monitoring. CAF and its metabolites, PARA, have been proposed as anthropogenic markers to assess the population size and trace the discharge of domestic wastewater in rivers and lakes [54]. Senta *et al*. reported the PARA loads in the wastewater reflected the population dynamics [25].We demonstrated the greatly improved correlation between PARA-normalized SARS-CoV-2 load per capita and the prevalence using a college town as an example (Fig. 11). Among 3 normalization scenarios (Fig. 11), only the PARA-normalized SARS-CoV-2 load per capita and PARA-normalized cases per capita yielded a statistically significant correlation (*rho =* 0.5878, p<0.05). Our results indicated that a fixed population often derived from population census is not ideal for long term monitoring. It can be challenging to capture the population dynamic during the COVID-19 pandemic with the conventional methodologies based on periodic public surveys (such as census taking), augmented with a wide array of demographic statistics. Most of the inaccurate population data often derived from aged or incomplete sources such as census surveys or utility customers billed (e.g., Anderson et al., 2004 [62]; Banta-Green *et al*., 2009 [63]; Clara *et al*., 2011[64]; Kasprzyk-Hordern *et al*., 2009 [65]; Neset *et al*., 2010 [66]; Ort *et al*., 2009 [67]; Rowsell *et al*., 2010 [68]; Tsuzuki, 2006[69]). Particularly during current pandemic, population dynamics often deviate significantly from the population estimated by the conventional methodologies due to the introduction of restrictions in control of the spread of SARS-CoV-2.

Unreliable population biomarkers often result in the poor correlation between the normalized SARS-CoV-2 levels and prevalence. For example, Feng *et al*. reported normalizing SARS-CoV-2 concentration in the wastewater to fecal marker HF183 and pMMoV reduced correlations in 5 and 8 of 12 WWTPs, respectively, compared to the correlation before normalization [45]. Greenwald *et al*. also reported normalizing SARS-CoV-2 load using crAssphage, pMMoV, and Bacteroides rRNA in the wastewater samples deteriorated the correlation with daily case number per capita in comparison with the correlation between non-normalized concentrations and daily case numbers [70]. According to our results, the worsen correlations could result from using fixed populations to normalize clinical cases.

## 5. CONCLUSION

Our findings suggested that the CAF metabolite, PARA, is a reliable population biomarker in SARS-CoV-2 wastewater-based epidemiology studies, due to its 1) better population indicators with higher accuracy, lower variability and higher temporal consistency as a population indicator to reflect the change in population dynamics and dilution in wastewater, 2) very limited exogenous sources, 3) high extraction efficiency with low variability in the extraction rates, 3) high stability, 4) resistance to chemicals in the wastewater, and 5) low sample volume requirement with simple sample preparation process. This chemical biomarker offers an excellent alternative to the currently CDC-recommended pMMoV genetic biomarker to help us understand the size, distribution, and dynamics of local populations for forecasting the prevalence of SARS-CoV2 within each sewershed. Furthermore, the regression functions embedded in the direct and indirect approaches of normalizing viral loads by biomarker could be applied to new data without known daily flow volume and population. Finally, the clinical cases should also be normalized by population dynamics when the correlation between SARS-CoV-2 and prevalence were examined. Based on the findings in this study, we recently launched a long-term study to compare the utility of CAF, PARA and pMMoV for SARS-CoV-2 population normalization cross 64 facilities in the Missouri.

## Data Availability

All data produced in the present study are available upon reasonable request to the authors.

## FUNDING AND ACKNOWLEDGEMENT

The authors would like to thank the Missouri Department of Health and Senior Services (DHSS) administrating the funding. We would like to express our gratitude to the Missouri Department of Natural Resources (DNR) for coordinating the sample collection. Research reported in this publication was supported by funding from the Centers for Disease Control and the National Institute on Drug Abuse of the National Institutes of Health under award number U01DA053893-01. We would also like to thank the Center for Agroforestry at University of Missouri, USDA/ARS Dale Bumpers Small Farm Research Center under agreement number 58-6020-6-001 from the USDA Agricultural Research Service for supporting part of this research. The content is solely the responsibility of the authors and does not necessarily represent the official views of the National Institutes of Health, the Centers for Disease Control or USDA-ARS.

## Notes

### Competing Interest Statement

The authors have declared no competing interest.

