## Supplementals for "Biomarkers Selection for Population Normalization in SARS-CoV-2 Wastewater-based Epidemiology"

**SUPPLEMENTAL**

Table S1. Site summary of Wastewater Treatment Plants and Samples Collected for Second Data Set in This Study

| No. | Project ID | City | Population Served | Source of Population | [a]Facility Capacity | Composite sampling mode | [b] Daily influent flow |
|---|---|---|---|---|---|---|---|
| 1 | CARTH | Carthage | 12,000 | Operator information | 7 | Time Based | 3.82 |
| 2 | ROLSE | Rolla | 15,000 | Operator information | 4.8 | Time Based | 4.39 |
| 3 | SDLCN | Sedalia | 7,500 | Operator information | 3.03 | Time Based | 3.03 |
| 4 | KCBLU | Kansas City | 287,500 | Operator information | 105 | Time Based | 101.47 |
| 5 | SFDSW | Springfield | 173,676 | Connections with population correction | 64 | Time Based | 48.56 |
| 6 | CASVL | Cassville | 3,300 | Operator information | 1.1 | Time Based | 1.01 |
| 7 | MONET | Monett | 9,100 | Operator information | 6 | Time Based | 4.40 |
| 8 | JOPTC | Joplin | 35,462 | Connections with population correction | 15 | Time Based | 16.10 |
| 9 | WARNE | Warrensberg | 7,990 | Operator information | 1.5 | Flow Based | 2.29 |
| 10 | CHARL | Charleston | 4,000 | Operator information | 1.5 | Time Based | 1.50 |
| 11 | ALBNY | Albany | 1,710 | Operator information | 0.49 | Time Based | 0.19 |
| 12 | MSDME | St. Louis | 451,367 | Operator information | 210 | Time Based | 97.40 |
| 13 | UNONW | Union | 7,378 | Connections with population correction | 1.5 | Time Based | 0.75 |
| 14 | FULTN | Fulton | 12,790 | Operator information | 2.9 | Time Based | 1.90 |
| 15 | MSDGG | Valley Park | 115,895 | Operator information | 21 | Time Based | 13.61 |
| 16 | SFDNW | Springfield | 26,078 | Connections with population correction | 6.8 | Time Based | 13.60 |
| 17 | MILAN | Milan | 1,960 | Operator information | 0.7 | Flow Based | 0.38 |
| 18 | HANBL | Hannibal | 16,000 | Operator information | 12 | Time Based | 4.14 |
| 19 | STPSC | St. Peters | 48,774 | Operator information | 9.5 | Time Based | 6.51 |
| 20 | JEFFC | Jefferson City | 75,000 | Operator information | 11 | Flow Based | 8.10 |
| 21 | NPSDS | FENTON | 44,035 | Connections with population correction | 4 | Time Based | 2.27 |
| 22 | JOPSC | Joplin | 11,800 | Connections with population correction | 7.2 | Time Based | 6.30 |
| 23 | SDLSE | Sedalia | 7,500 | Connections with population correction | 2.6 | Time Based | 4.29 |
| 24 | FRMTN | Farmington | 9,525 | Operator information | 2.75 | Time Based | 2.87 |
| 25 | WARNW | Warrensburg | 6,235 | Operator information | 1.5 | Flow Based | 2.20 |
| 26 | SDLNO | Sedalia | 8,250 | Operator information | 2.5 | Time Based | 1.47 |
| 27 | CAROL | Carrollton | 3,784 | Operator information | 1.5 | Time Based | 0.73 |
| 28 | MACON | Macon | 5,471 | Operator information | 2.5 | Time Based | 1.12 |
| 29 | SKSTN | Sikeston | 17,000 | Operator information | 5 | Time Based | 2.40 |
| 30 | MSDBP | St. Louis | 306,647 | Operator information | 150 | Time Based | 93.80 |

| | | | | | | | |
|---|---|---|---|---|---|---|---|
| 31 | STROB | St. Roberts | 4,085 | Operator information | 1 | Time Based | 0.64 |
| 32 | PRYVL | Perryville | 9,000 | Operator information | 1.8 | Time Based | 1.09 |
| 33 | DEXTW | Dexter | 2,400 | Operator information | 0.78 | Time Based | 0.75 |
| 34 | COLMB | Columbia | 123,180 | Operator information | 20.6 | Time Based | 15.70 |
| 35 | MSDLM | St. Louis | 66,738 | Operator information | 15 | Time Based | 9.55 |
| 36 | MSDFN | St. Louis | 24,174 | Operator information | 6.75 | Time Based | 4.05 |
| 37 | MRSHL | Marshall | 13,000 | Operator information | 7.1 | Time Based | 2.38 |
| 38 | STJOE | St. Joseph | 58,200 | Connections with population correction | 27 | Time Based | 17.73 |
| 39 | MSDMR | St. Louis | 174,537 | Operator information | 38 | Time Based | 17.70 |
| 40 | LBVAT | Independence | 360,000 | Population Equivalent | 52 | Time Based | 85.66 |
| 41 | BROOK | Brookfield | 4,600 | Operator information | 2 | Time Based | 1.12 |
| 42 | CAPEG | Cape Girardeau | 38,000 | Operator information | 11 | Flow Based | 7.64 |
| 43 | MEXCO | Mexico | 11,500 | Operator information | 3 | Time Based | 3.40 |
| 44 | MARSH | Marshfield | 8,000 | Operator information | 1.5 | Time Based | 2.25 |
| 45 | NIXAF | Nixa | 20,000 | Operator information | 4 | Time Based | 2.00 |
| 46 | NEVAD | Nevada | 3,674 | Connections with population correction | 2 | Time Based | 2.75 |
| 47 | WARSW | Warsaw | 2,204 | Connections with population correction | 0.45 | Flow Based | 0.70 |
| 48 | BLIVR | Bolivar | 10,500 | Operator information | 2.55 | Time Based | 1.37 |
| 49 | ELDON | Eldon | 4,895 | Operator information | 1 | Time Based | 0.98 |
| 50 | WPLAN | West Plains | 12,000 | Operator information | 3 | Time Based | 5.42 |
| 51 | PACIF | Pacific | 7,001 | Operator information | 2 | Time Based | 1.24 |
| 52 | LIBTY | Liberty | 35,300 | Operator information | 5 | Time Based | 4.60 |
| 53 | KCWST | Kansas City | 50,000 | Connections with population correction | 22.5 | Time Based | 18.13 |
| 54 | KCBIR | Kansas City | 55,000 | Connections with population correction | 20 | Time Based | 15.10 |
| 55 | KCROB | Kansas City | 12,000 | Operator information | 2.8 | Time Based | 3.19 |
| 56 | KCFSR | Kansas City | 12,000 | Operator information | 2 | Time Based | 1.17 |
| 57 | KCTDC | Kansas City | 9,000 | Operator information | 3.4 | Time Based | 3.13 |
| 58 | WLOSP | Willow Springs | 2,100 | Operator information | 0.4 | Time Based | 0.81 |
| 59 | MEMPH | Memphis | 1,822 | Operator information | 0.21 | Time Based | 0.42 |
| 60 | WASHN | Washington | 15,000 | Operator information | 4 | Time Based | 2.26 |
| 61 | Anonymous facility #1 | - | 10,559 | Operator information | 5.3 | Time Based | 2.73 |
| 62 | Anonymous facility #2 | - | 6,155 | Operator information | 3.4 | Time Based | 1.59 |

| | | | | | | | |
|---|---|---|---|---|---|---|---|
| 63 | Anonymous facility #3 | - | 12,000 | Operator information | 9 | Time Based | 2.80 |
| 64 | Anonymous facility #4 | - | 900 | Operator information | 0.14 | Time Based | 0.06 |

[a] Unit: million gallon per day (MGD).

[b] Samples were collected during the week of May 10th, unit: MGD.

Table S2. Site summary of Wastewater Treatment Plants and Samples Collected for Third Data Set in This Study

| No. | Project ID | City | Population Served | Source of Population | [a] Facility Capacity | Composite sampling mode |
|---|---|---|---|---|---|---|
| 1 | College town | Columbia | 123,180 | Operator information | 20.6 | Time Based |
| 2 | Tourist town | Anonymous facility #1 | 10,559 | Operator information | 5.3 | Time Based |

[a] Unit: million gallon per day (MGD).

Table S3. Results of correlations between log-transformed biomarker concentration and population concentration in wastewater.

| Biomarker | No data transformation | | Log transformation | |
|---|---|---|---|---|
| | Correlation (*r*) | Regression function | Correlation (*r*) | Regression function |
| **CAF** | 0.81 | y = 0.0066x + 7.1668 | 0.88 | y = 1.0577x - 2.3383 |
| **PARA** | 0.77 | y = 0.0015x + 4.3207 | 0.86 | y = 0.8819x - 2.2076 |
| **5-HIAA** | 0.59 | y = 0.0005x + 0.9342 | 0.72 | y = 1.2553x - 4.1942 |
| **pMMoV** | 0.60 | y = 166.29x - 45396 | 0.70 | y = 1.0364x + 1.996 |
| **CRE** | 0.06 | y = 0.0026x + 87.104 | 0.06 | y = 0.0026x + 87.104 |

Table S4. Results of correlations between log-transformed biomarker load and population contributing to wastewater.

| Biomarker | No data transformation | | Log transformation | |
|---|---|---|---|---|
| | Correlation (*r*) | Regression function | Correlation (*r*) | Regression function |
| **CAF** | 0.99 | y = 0.0215x + 209.66 | 0.97 | y = 0.9807x - 1.4619 |
| **PARA** | 0.97 | y = 0.0061x + 75.589 | 0.97 | y = 0.9993x - 2.0667 |
| **5-HIAA** | 0.98 | y = 3E+11x - 2E+15 | 0.87 | y = 1.0404x - 2.8396 |
| **pMMoV** | 0.98 | y = 0.0027x + 1.1872 | 0.92 | y = 1.0889x + 10.87 |
| **CRE** | 0.22 | y = 0.0042x + 1166.6 | 0.33 | y = 0.5842x + 0.1257 |

Table S5. Statistical Summary of $C_1$ to $C_0$ Fold Changes from Selected Biomarker.

| Biomarkers | Count | Sum | Average | Standard deviation |
|---|---|---|---|---|
| **CAF** | 24 | 24.994 | 1.041 | 0.311 |
| **PARA** | 24 | 25.364 | 1.057 | 0.389 |
| **pMMoV** | 24 | 27.972 | 1.166 | 0.737 |
| **5-HIAA** | 24 | 27.598 | 1.150 | 0.661 |

Table S6. Statistical Summary of $C_2$ to $C_0$ Fold Changes from Selected Biomarker.

| Biomarkers | Count | Sum | Average | Standard deviation |
|---|---|---|---|---|
| **CAF** | 24 | 23.204 | 0.967 | 0.324 |
| **PARA** | 24 | 25.018 | 1.042 | 0.341 |
| **pMMoV** | 24 | 24.064 | 1.003 | 0.586 |
| **5-HIAA** | 24 | 35.292 | 1.470 | 1.144 |
| **CRE** | 24 | 138.000 | 5.750 | 17.343 |

Table S7. Statistical Summary of $C_1$ to $C_0$ Fold Changes from CAF and PARA.

| Biomarkers | Count | Sum | Average | Standard deviation |
|---|---|---|---|---|
| **CAF** | 64 | 61.459 | 0.960 | 0.446 |
| **PARA** | 64 | 61.266 | 0.957 | 0.498 |

Table S8. Statistical Summary of $C_2$ to $C_0$ Fold Changes from CAR and PARA.

| Biomarkers | Count | Sum | Average | Standard deviation |
|---|---|---|---|---|
| **CAF** | 64 | 57.151 | 0.893 | 0.433 |
| **PARA** | 64 | 59.952 | 0.937 | 0.428 |

Table S9. Studies reporting correlation between SARS-CoV-2 and prevalence data.

| Refence | sample | SARS-CoV-2 normalization and unit | Clinical case scenario | Correlation between case and viral load | Correlation analysis |
|---|---|---|---|---|---|
| Green *et al.* 2020. [1] | Wastewater | crAssphage<br>(The ratio of SARS-CoV-2:crAssphage) | cumulative incidence of COVID-19 | The ratio of SARS-CoV-2:crAssphage visually correlates with the cumulative incidence of COVID-19. | - |
| Medema *et al.* 2020 [2] | Wastewater | -<br>(Concentration of SARS-CoV-2 RNA) | The cumulative number of reported COVID-19 cases<br>(Reported COVID-19 cases/100K people) | $R^2$ = 0.59-0.79 | Person |
| Peccia *et al.*. 2020 [3] | Sludge | Volume of sludge<br>(Viral copies per sludge) | Daily Number of positive SARS-CoV-2 test results, Number of COVID-19 admissions<br>(Case number/day) | SARS-CoV-2 RNA concentrations in sludge were 0–2 d prior to SARS-CoV-2 positive test results by date of specimen collection, 1–4 d ahead of local hospital admissions and 6–8 d ahead of SARS-CoV-2 positive test results by reporting date. | - |
| Gonzalez *et al.* 2020 [4] | Wastewater | Population<br>(copies/person) | (Confirmed case/city population) | Fluctuations in population normalized loading rates in several of the WWTP service areas agreed with known outbreaks during the study. | - |
| Weidhaas *et al.* 2021 [5] | Wastewater | Population<br>(Million viral GC/capita/day) | Weekly new cases<br>(Weekly case rate/100K people) | *Rho* = 0.54 for all facility,<br>*rho* = 0.82 for LCCWWTP and *rho* = 0.96 for HCWWTP. | Spearman's |
| Westhaus *et al.* 2021[6] | Wastewater | -<br>(Copies per day) | The cumulative number of reported COVID-19 cases<br>(Case number) | SARS- CoV-2 concentration in wastewater and prevalence data were not significantly correlated. | - |
| D'Aoust *et al.* 2021 [7] | Influent post grit solids (PGS) and primary clarified sludge (PCS) | pMMoV<br>(copies/copies of pMMoV) | Daily number of tests performed and the number of active COVID-19 cases<br>(cases/100K population) | R = 0.498 for N1,<br>R = 0.639 for N2<br>in Ottawa | Pearson |

| | | | | | |
|---|---|---|---|---|---|
| Feng *et al.* 2021 [8] | Wastewater | Population, pMMoV, BCoV, HF183 (Copies/L/person or copies/copies biomarker) | 7 day moving average of diagnosed COVID-19 cases (cases/100K people) | *Rho* was 0.73 and 0.74 for raw concentrations and concentrations adjusted per capita respectively. Normalizing to HF183 and pMMoV reduced correlations in 5 and 8 of 12 WWTPs respectively. | Spearman's |
| Greenwald *et al.* 2021 [9] | Wastewater | crAssphage, pMMoV, Bacteroides rRNA (N1 copies/copies of biomarker) | Moving average of daily per capita new cases (Case number/person) | Kendall tau-b=0.43 for N1 concentration vs case number/person.<br>Kendall tau-b=0.38 for N1/pMMoV.<br>Kendall tau-b=0.35 for N1/Bacteroides rRNA. | Kendall's Tau-b coefficients |
| Ai *et al.* (2021) [10] | Wastewater | pMMoV<br>crAssphage<br>(N2 Copies/copies of biomarker) | average daily case numbers (Case number) | Rho= 0.78 for N2 concentration<br>Rho=0.79 for pMMoV-normalized N2 concentration<br>Rho= 0.55 for crAssphage-normalized N2 concentration | Spearman's |
| Wolfe *et al.* (2021) [11] | Primary settled solids | pMMoV<br>(N1 or N2 Copies/copies of biomarker) | 7-d smoothed new COVID-19 cases in the 142 sewershed the day the wastewater sample was collected<br>(Case number) | N1 and/or N2, N1 and/or N2 normalized by PMMoV, and N1 and/or N2 scaled by the LHS of eq 3 were positively associated with COVID-19 cases at all seven POTWs ($p < 0.05$).<br>Normalizing or scaling does not consistently improve correlations with COVID-19 cases within sewershed | Kendall's tau |
